# Treatment Preferences For Comorbid Obesity and Obstructive Sleep Apnea (PRO-CON OSA) Survey: Patient and Provider Preferences for CPAP and/or Tirzepatide

**DOI:** 10.1101/2025.10.02.25337176

**Authors:** Christopher N. Schmickl, Athiwat Tripipitsiriwat, Babak Mokhlesi, Monica Mallampalli, Brandon Nokes, Vaishnavi Kundel, Kathy Page, Christina Finch, Lucas Donovan, Mira Tadros, Ravi S. Aysola, Andrey Zinchuk, Tracy Zvenyach, M. Safwan Badr, Sanjay R. Patel, Jeremy E. Orr, Robert L. Owens, Chris Lindsell, Jennifer L. Martin, Atul Malhotra, the PARADIGM-SLEEP Investigators (see Acknowledgements)

## Abstract

**Purpose:** Approximately one of ten US adults has comorbid obesity and obstructive sleep apnea (COBOSA). Traditionally, sleep medicine management of COBOSA focused on continuous positive airway pressure (CPAP). Recently, tirzepatide (a once-weekly injection) was approved for COBOSA after demonstrating substantial reduction of weight and OSA severity in efficacy trials. We assessed patient and provider attitudes towards these therapies for COBOSA.

**Methods:** We conducted an online survey (November 2024–August 2025) targeting US adults with OSA and/or obesity (“patients”) and sleep medicine providers. The survey assessed treatment acceptability, preferences, and informational needs.

**Results:** The main analysis included 461 patients (86% with sleep apnea diagnosis, 49% with COBOSA) and 114 providers. Overall, 70% of respondents found both CPAP and tirzepatide at least somewhat acceptable, with significantly different response patterns (P<.001): providers found CPAP more acceptable than tirzepatide, whereas patients rated both therapies similar. When asked to choose a preferred long-term therapy assuming equal effectiveness, patients favored tirzepatide (48% vs 21%), while providers preferred CPAP (52% vs 27%). Providers with experience prescribing injectable weight-loss medications were more aligned with patient views. Both groups supported combination therapy, though patients were less enthusiastic than providers (61% vs 86%). Both groups valued a wide range of outcomes for decision making—across symptom, sleep, and cardiometabolic health domains—and emphasized the importance of safety, long-term data, and costs.

**Conclusions:** Patients and providers view CPAP and/or tirzepatide as acceptable options for COBOSA, but preferences diverge. Given equipoise, comparative effectiveness trials are urgently needed to guide individualized treatment strategies.

**BRIEF SUMMARY:** *Current Knowledge/Study Rationale:* The weight-loss medication tirzepatide has recently emerged as a potential treatment option for patients with obstructive sleep apnea (OSA) and comorbid obesity (COBOSA), but little is known about patient and provider attitudes toward this therapy compared with the current first-line treatment of continuous positive airway pressure (CPAP) or what information they need to make informed choices between these options.

*Study Impact:* This survey demonstrates high levels of acceptability for both tirzepatide and CPAP, supporting clinical equipoise. It also highlights the priorities, concerns, and decision-making factors most relevant to patients and providers, providing a foundation for future comparative trials and implementation efforts that are aligned with stakeholder needs and preferences.

## INTRODUCTION

The advent of highly effective and well-tolerated weight loss medications is reshaping clinical practice across multiple specialties, including sleep medicine.^1^ Both obesity and obstructive sleep apnea (OSA) are increasingly common and frequently co-occur. For example, 40% of US adults have obesity, and an estimated 26% have OSA—of which 40–60% of cases are attributable to excess weight.^2–8^ This overlap defines a clinically important, highly prevalent subgroup: <u>c</u>omorbid <u>ob</u>esity and <u>OSA</u> (COBOSA), which affects roughly 1 in 10 US adults and is associated with a wide range of symptoms and cardiometabolic complications.^9, 10^

OSA is characterized by repetitive upper airway collapse during sleep, resulting in intermittent hypoxemia and sleep fragmentation. Untreated OSA may contribute to neurocognitive impairment (e.g., excessive sleepiness,^11^memory impairment^12^) and is associated with adverse cardiovascular outcomes, including hypertension,^13^ coronary artery disease,^14^ and increased mortality.^15^Continuous positive airway pressure (CPAP) therapy remains the first-line treatment for symptomatic OSA and is highly efficacious in preventing upper airway collapse.^9, 16–18^ However, adherence is suboptimal, with average nightly use around 3–5 hours,^19, 20^ limiting its real-world effectiveness.^20, 21^

Although excess weight is a major contributor to OSA, weight loss has traditionally been viewed as a secondary goal since substantial and sustained weight reduction has been difficult to achieve. The American Thoracic Society (2018) clinical practice guidelines recommend comprehensive lifestyle interventions be incorporated into the routine managegement of patients with OSA and comorbid overweight/obesity.^22^ However, access to comprehensive lifestyle programs is often limited, and even in ideal settings, weight loss is temporary and modest (typically 2-5%).^23–26^ Prior anti-obesity medications (e.g., phentermine-topiramate, naltrexone-bupropion) yielded limited weight loss (5-10%) and variable tolerance.^10, 27, 28^ Thus, OSA guidelines either ignored^18, 29^ or only weakly endorsed^22^ pharmacologic weight loss strategies. As a result, a CPAP-centric paradigm has dominated OSA management for more than three decades.

Tirzepatide, a once-weekly self-injected Glucagon-Like Peptide-1/Glucose-Dependent Insulinotropic Polypeptide (GLP-1/GIP) receptor agonist, has recently emerged as a promising treatment for obesity and related conditions. In the SURMOUNT trials,^24, 25, 30^ one year of tirzepatide led to ∼20% weight loss, which in the SURMOUNT-OSA study^25^ translated to a ∼55% reduction in OSA severity. This magnitude is comparable to the real-world effectiveness of CPAP when accounting for typical treatment adherence (i.e the effective AHI). In trial settings, tirzepatide also improved several OSA-related symptoms, blood pressure, and other cardiometabolic risk factors beyond what is typically achieved with CPAP alone (**E-Table 1**).^25, 31^ SURMOUNT-OSA also reaffirmed a favorable safety profile, with mostly mild-to-moderate, transient gastrointestinal side effects, leading to tirzepatide discontinuation in <5% of patients.^24, 25^ Based on these results, the US Food and Drug Administration approved tirzepatide on December 20, 2024, for the treatment of COBOSA.^32^

**Table 1.**
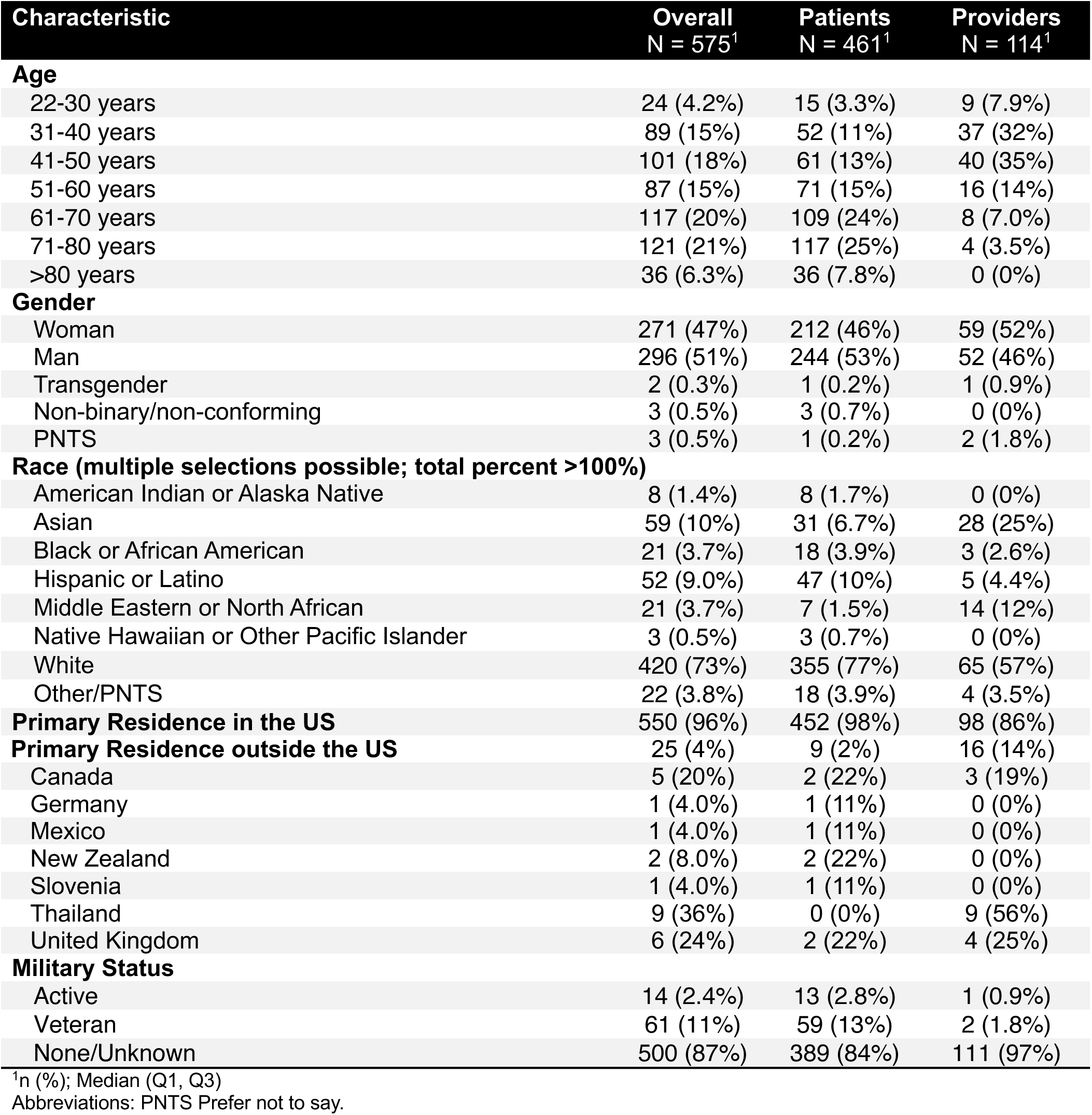
General Characteristics of all Included Survey Respondents.

In the absence of comparative effectiveness trials, treatment decisions for COBOSA—whether to use CPAP, tirzepatide, or both—are currently guided by individual preferences and expert opinion.^33, 34^ To inform comparative effectiveness research, future guidelines, and shared decision-making in clinical practice, we sought to conduct a national survey to assess patient and provider attitudes toward CPAP and/or tirzepatide for COBOSA, and identify key informational needs for their decision making.

## METHODS

### Survey Development

Using REDCap, we developed an open online survey in October 2024. The landing page outlined the survey’s purpose, eligibility criteria, estimated completion time, types of information queried, and emphasized that participation was voluntary and anonymous, with completion implying consent.

Information about the principal investigator (CNS) and the overseeing institutional review board (IRB) were provided as well. Participants were then asked to identify themselves as a sleep provider (MD, NP, etc) managing OSA (hereafter “provider”) or not (hereafter “patient”).

Three sequential forms (i.e., web pages) then assessed participants’:

1) General characteristics and experiences (up to 35 items for patients and 12 items for providers, with branching logic),
2) Acceptability and preferences for CPAP and/or tirzepatide for treating COBOSA (4 items, which followed after a brief and balanced overview of COBOSA and treatments **E-Figure 1**),
3) Perceived importance of various outcomes and other factors for decision making (up to 28 items for patients and 25 items for providers).

**Figure 1.**
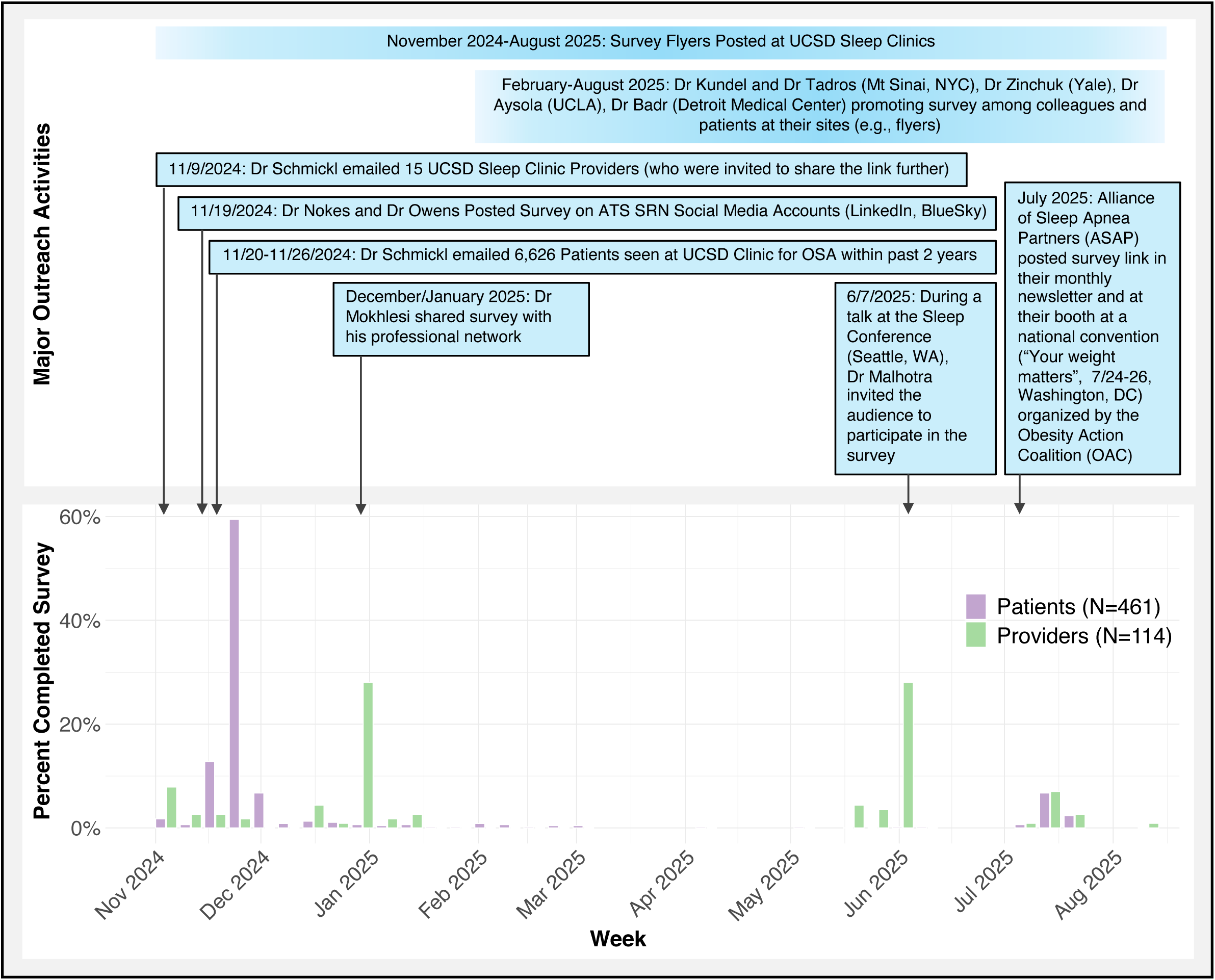
Timeline of Outreach Activities and Survey Completions.

Branching logic ensured that providers answered questions from the perspective of writing prescriptions, while patients responded as potential users. The survey draft was created by a physician board-certified in both sleep and obesity medicine (CNS), and pilot tested by a physician-scientist with experiences in sleep and patient-reported outcomes research (LD), a pulmonary/sleep medicine physician (AT), and a clinical sleep psychologist (JM). Further refinements were made based on cognitive interviews with patient and clinician partners (KP, CF).

To complete a form all questions had to be answered, and nearly all questions included an “unsure” and/or “prefer not to say” option. Respondents could pause and return later using a code, but could not revise responses from earlier forms.

### Survey Distribution

We recruited a convenience sample, targeting adults with OSA and/or obesity, as well as sleep medicine providers across the United States. In order to capture most potential future stakeholders, the only eligibility criterion was age 21 years or above, regardless of comorbidities.

Recruitment began in November 2024 with flyers (**E-Figures 2 & 3**) that were posted at the UCSD sleep clinic and emailed to 6,626 patients seen there for OSA during the preceding 2 years. Flyers were also shared with colleagues and national partner organizations (e.g., American Thoracic Society Sleep Respiratory & Neurobiology [ATS-SRN] section, Alliance of Sleep Apnea Partners [ASAP], Obesity Action Coalition [OAC]) who promoted the survey locally and online. **Figure 1** summarizes major outreach activities and resulting responses.

**Figure 2.**
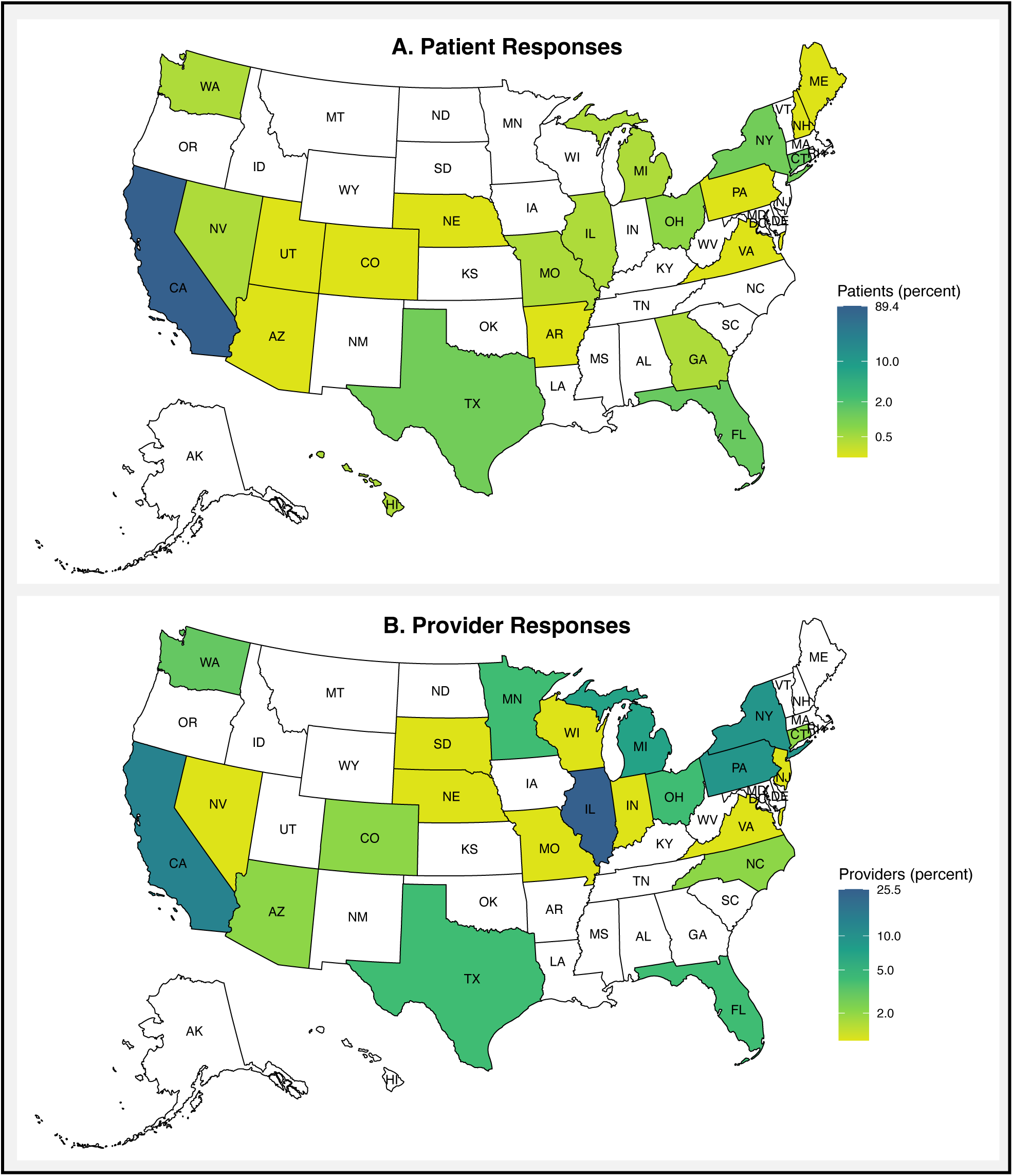
US Survey Responses by State. 98% (452) of participating patients and 86% (98) of providers reported their primary residence to be in the United States. Among those, patient responses were concentrated in California (**Panel A**), whereas provider responses were more broadly distributed across US regions (**Panel B**).

The survey was open from November 9, 2024 to August 11, 2025, with most responses received between November 2024 and January 2025. Responses were anonymous and uncompensated. While participants were asked to complete the survey only once, anonymity precluded detection of potential duplicate entries.

The UCSD IRB determined that this study (#811561) met the criteria for exempt research under 45 CFR 46.104(d) 2 (i), and granted a partial waiver to use protected health information to email invitations to select UCSD sleep clinic patients (see above).

### Analysis and Reporting

Data were summarized as number (percent) or median (interquartile range), as appropriate. Response patterns between patients and providers for categorical variables were compared using Fisher’s exact tests. For free-text responses, we used structural topic modeling (STM)^35^ and large language models (ChatGPT 40, OpenAI) in combination with manual reviews to identify common themes.

All analyses were performed in R (version 4.4.1; major packages: usmap, stm), using P<.05 to denote statistical significance. The study adhered to the Checklist for Reporting Results of Internet E-Surveys (CHERRIES).^36^

## RESULTS

The survey was started by 677 individuals, of whom 575 (85%) completed the treatment acceptability and preference form and were included in the main analysis (461 patients, 114 providers, **E-Figure 4**).

### Characteristics of Survey Participants

Overall, 47% of participants identified as women, 27% as non-white, and 9% as Hispanic. Providers were more racially diverse than patients (43% vs 23% non-white) and generally younger (75% under age 50 vs most patients >50 years, **Table 1**). Most patients (98%) and providers (86%) resided in the US, with patient responses concentrated in California, while provider responses were more geographically dispersed (**Figure 2**).

Among the 461 patients, 87% had overweight or obesity, and 86% reported a diagnosis of OSA. Nearly half (49%) met criteria for COBOSA. Most reported at least one comorbidity such as hypertension, and frequent symptoms including sleepiness, fatigue or unrefreshing sleep. The majority (73%) had attempted weight loss three or more times, 97 (21%) had used GLP-1 receptor agonists like tirzepatide, and 355 (77%) had used CPAP (**Table 2**).

**Table 2.**
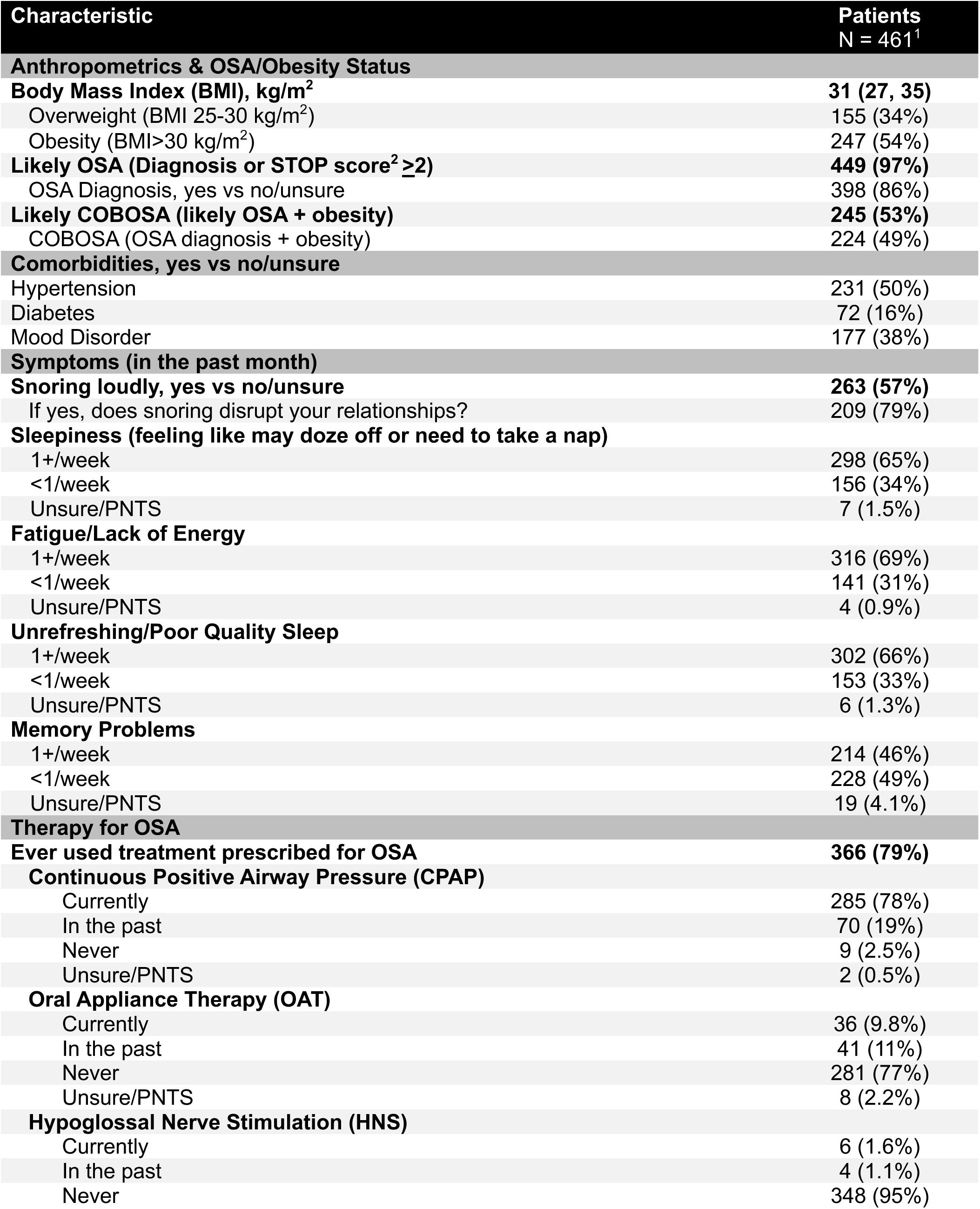
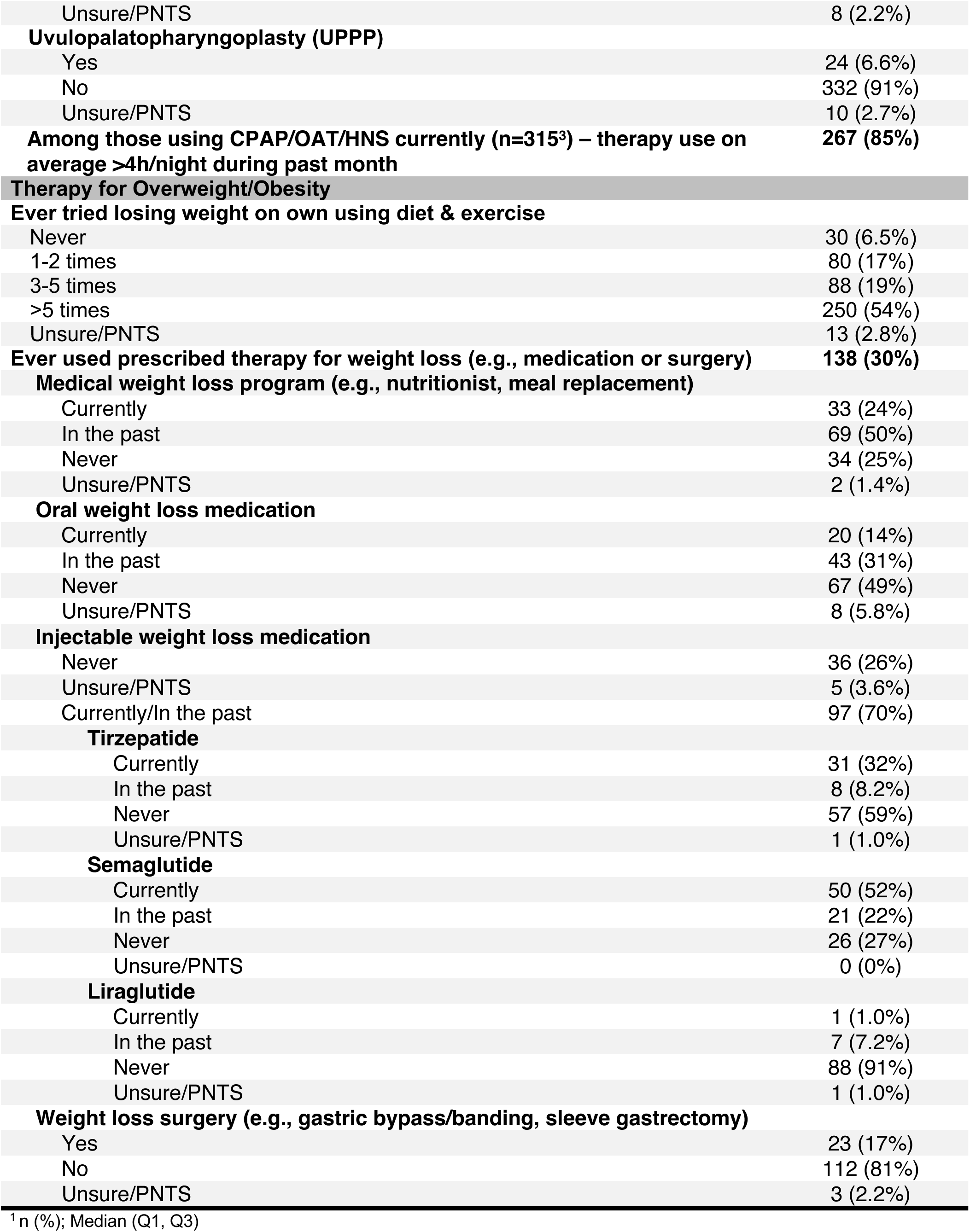
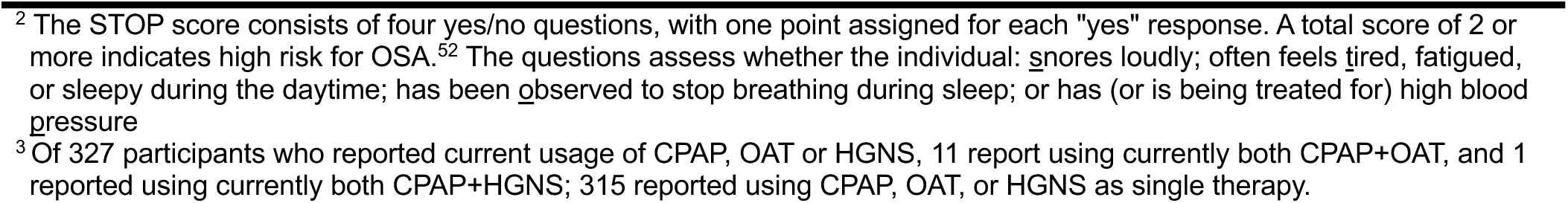
Patients Only: Clinical Characteristics and History.

The 114 providers practiced sleep medicine in a variety of settings, including academic (51%), community (28%), private practice (22%) and Veterans Affairs (8%) clinics. Nearly half (46%) had over 10 years of experience treating OSA, and 52% managed 30+ patients with OSA per week. Estimates of COBOSA prevalence varied, but 61% believed it affected at least half their patients; the most common estimate (24%) was 61-70%. While 94% reported extensive experience prescribing CPAP, only 11% had “a lot” of experience with injectable weight loss medications, and 47% reported none (**Table 3**).

**Table 3.**
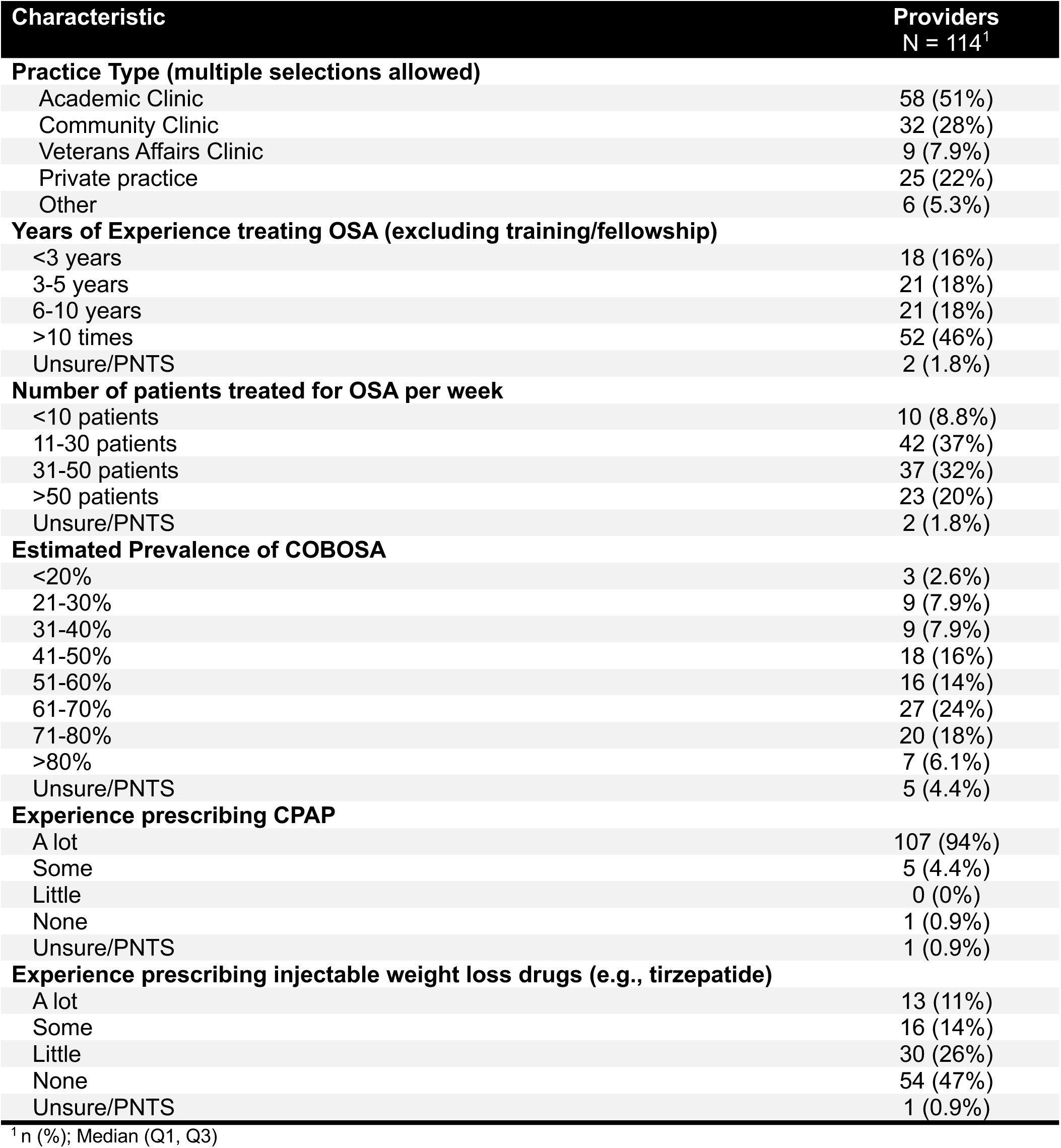
Providers only: Practice Setting and Experiences.

| **Characteristic** | **Providers**<br>**N = 114<sup>1</sup>** |
| --- | --- |
| **Practice Type (multiple selections allowed)** |  |
| Academic Clinic | 58 (51%) |
| Community Clinic | 32 (28%) |
| Veterans Affairs Clinic | 9 (7.9%) |
| Private practice | 25 (22%) |
| Other | 6 (5.3%) |
| **Years of Experience treating OSA (excluding training/fellowship)** |  |
| <3 years | 18 (16%) |
| 3-5 years | 21 (18%) |
| 6-10 years | 21 (18%) |
| >10 times | 52 (46%) |
| Unsure/PNTS | 2 (1.8%) |
| **Number of patients treated for OSA per week** |  |
| <10 patients | 10 (8.8%) |
| 11-30 patients | 42 (37%) |
| 31-50 patients | 37 (32%) |
| >50 patients | 23 (20%) |
| Unsure/PNTS | 2 (1.8%) |
| **Estimated Prevalence of COBOSA** |  |
| <20% | 3 (2.6%) |
| 21-30% | 9 (7.9%) |
| 31-40% | 9 (7.9%) |
| 41-50% | 18 (16%) |
| 51-60% | 16 (14%) |
| 61-70% | 27 (24%) |
| 71-80% | 20 (18%) |
| >80% | 7 (6.1%) |
| Unsure/PNTS | 5 (4.4%) |
| **Experience prescribing CPAP** |  |
| A lot | 107 (94%) |
| Some | 5 (4.4%) |
| Little | 0 (0%) |
| None | 1 (0.9%) |
| Unsure/PNTS | 1 (0.9%) |
| **Experience prescribing injectable weight loss drugs (e.g., tirzepatide)** |  |
| A lot | 13 (11%) |
| Some | 16 (14%) |
| Little | 30 (26%) |
| None | 54 (47%) |
| Unsure/PNTS | 1 (0.9%) |

<sup>1</sup> n (%); Median (Q1, Q3)

### Treatment Acceptability and Preferences

Over 70% of all respondents rated both CPAP and tirzepatide as at least “somewhat” acceptable for newly diagnosed COBOSA, though response patterns differed significantly between patients and providers (P <.001). Providers were more likely to rate CPAP as “very” acceptable compared to patients (95% vs 59%, **Figure 3A**). For tirzepatide, patients reported similar acceptability as for CPAP, and provider responses were more aligned with patient views for this treatment (**Figure 3B**).

**Figure 3.**
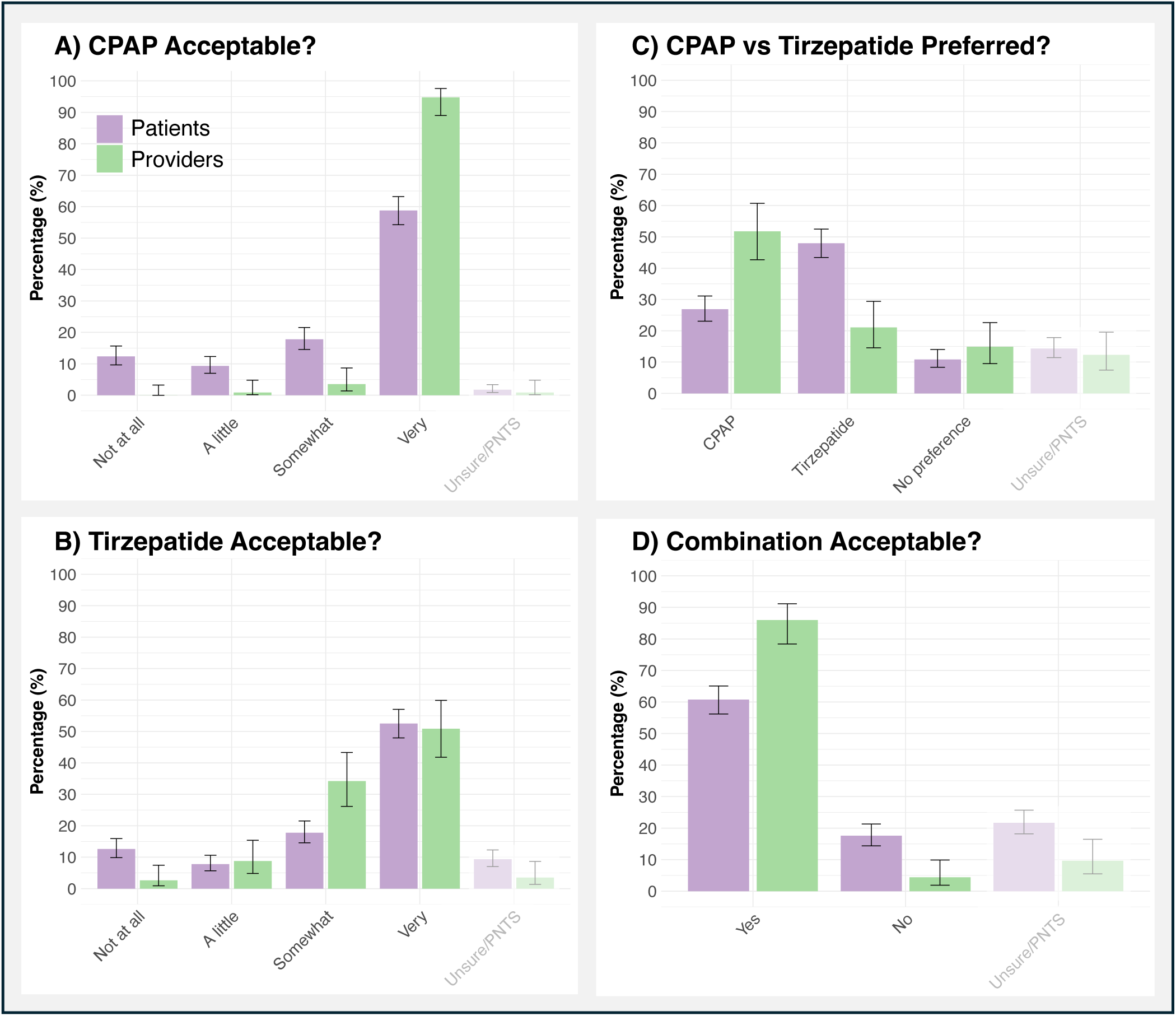
Treatment Acceptability and Preferences. If you had (a patient with) newly diagnosed, untreated COBOSA, how acceptable would you find it to use/prescribe CPAP (**Panel A)** or tirzepatide **(Panel B**). If there was strong evidence that both Tirzepatide and CPAP treat sleep apnea and associated risks/ symptoms similarly well, which one would you prefer long-term? (**Panel C**). If there was strong evidence that combining both CPAP + Tirzepatide leads to greater improvements of sleep apnea and associated risks/symptoms than using either CPAP or tirzepatide alone, would you be willing to use/prescribe both of them together long-term? (**Panel D**). Provider responses are green, non-provider (i.e., patients) responses are purple. Response patterns differed significantly between patients and providers across all four assessments (Fisher’s exact test, P<.001). Bars reflect 95% confidence intervals. PNTS = Prefer not to say.

**Figure 4.**
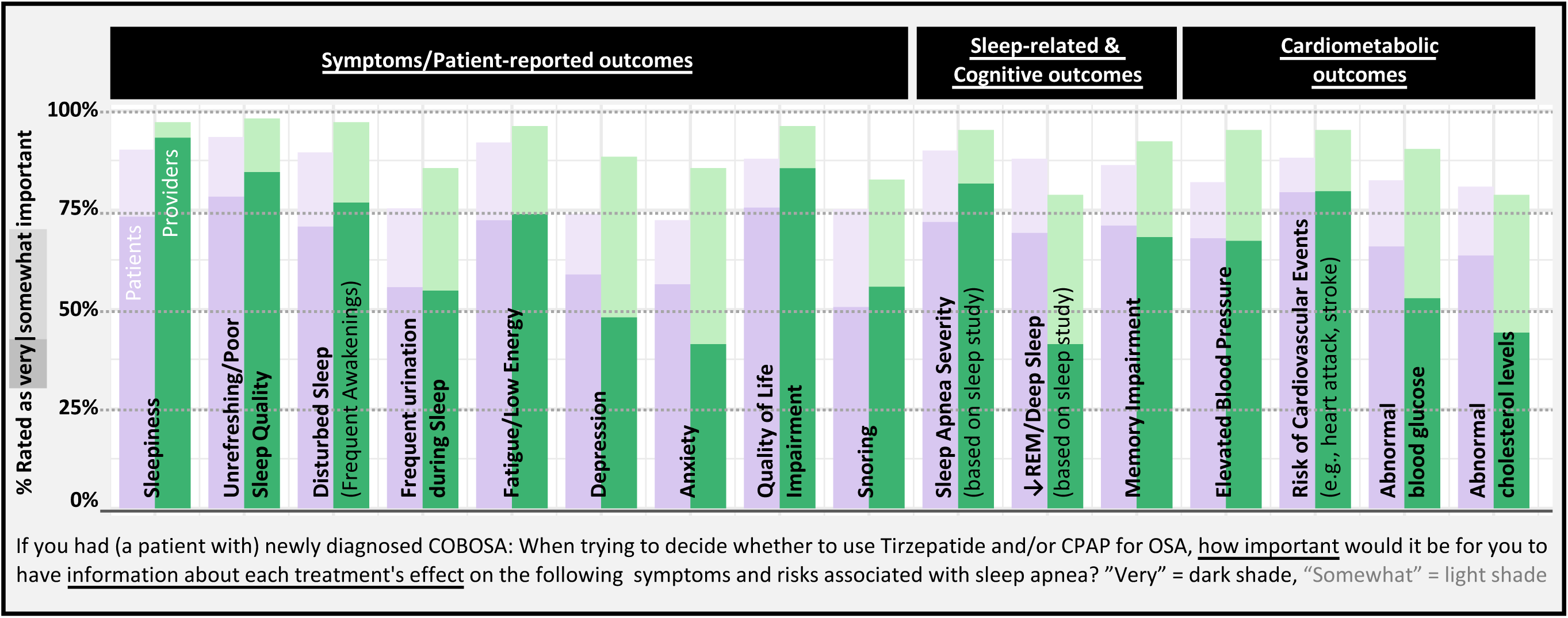
Information Patients (n=440) and Providers (n=104) Need for Decision Making.

When asked to choose a preferred long-term treatment—assuming both options were equally effective—patients favored tirzepatide (48% vs 21%), while providers preferred CPAP (52% vs 27%, **Figure 3C**). However, among the 29 providers reporting at least “some” experience using injectable weight-loss medications, preferences shifted towards patients’ views, with 41% favoring tirzepatide (**E-Figure 5**).

Both groups supported combination therapy if it offered superior benefits, though patients were less enthusiastic than providers (61% vs 86% combination acceptable; **Figure 3D**).

### Information Needed for Decision-Making

A total of 544 participants (440 patients and 104 providers) completed the outcomes form. Both groups expressed a need for information about the effects of both CPAP and tirzepatide across a broad range of outcomes. All 16 outcomes presented—spanning three key decision-driving domains: symptoms, sleep measures, and cardiometabolic health—were rated as at least “somewhat” important by over 72% of respondents, with more than 41% rating each outcome as “very” important (**Figure 4**).

Descriptively, the highest priority outcomes included sleepiness, sleep quality, quality of life, OSA severity, and cardiovascular risk. Patient and provider ratings were largely aligned; however, providers appeared to place slightly more emphasis on sleepiness, whereas patients seemed to attribute relatively more importance to improvements in sleep architecture.

When asked about eight potential factors that could influence treatment decisions—side effects, costs, ease of regular use, effect on weight, and time course of effect (for all respondents); ease of obtaining a prescription (patients only); and prescriber support and training (providers only)—over 83% of respondents rated each factor as at least “somewhat” important, with more than 58% rating each as “very” important. Patient and provider ratings were generally consistent (**E-Figure 6**).

Additionally, 118 participants provided free-text comments when asked whether any other information or considerations would be important for their treatment decisions. All respondents emphasized the importance of safety, long-term data, and costs/insurance coverage (**Table 4**). Among patients, other important themes included drug interactions, effects on comorbidities, and logistical barriers. For providers, over 10% of comments related to clinical monitoring and follow-up, administrative and staffing burden, comparative effectiveness, and consequences of therapy discontinuations (see **E-Tables 2 & 3** for representative quotes).

**Table 4.** Other Information Needed, and Other Important Factors for Decision Making. Some comments related to more than one theme.

| Themes | 92 Patients<sup>1</sup><br>(n = 132 themes) | 26 Providers<sup>1</sup><br>(n = 48 themes) |
| --- | --- | --- |
| **Clinical Considerations** |  |  |
| Safety Data | 27% (36) | 13% (6) |
| Long-term Data | 8% (10) | 6% (3) |
| Drug Interactions | 7% (9) | 2% (1) |
| Effect on Comorbidities | 9% (12) | 2% (1) |
| Special Populations | 3% (4) | 4% (2) |
| Effect/Management after Discontinuation | 5% (7) | 10% (5) |
| Comparative Effectiveness | 5% (7) | 10% (5) |
| **Evidence & Mechanism** |  |  |
| Mechanisms of Action | 5% (7) | 0% (0) |
| Quality of Evidence | 5% (7) | 0% (0) |
| Trust/Confidence | 5% (7) | 0% (0) |
| **Systems Issues, Logistics, and Other** |  |  |
| Costs and Insurance Coverage | 8% (11) | 10% (5) |
| Logistical Barriers | 6% (8) | 0% (0) |
| Clinical Monitoring, Protocols, & F/u Requirements | NA | 17% (8) |
| Administrative and Staffing Burden/Support | NA | 19% (9) |
| Other | 5% (7) | 6% (3) |

<sup>1</sup> Values shown as % (n); NA = not applicable.

### Impact of Nutrition Counselling Requirements

When participants were asked how a mandatory 1-hour, in-person dietary counseling visit each month for six months would affect their likelihood of using a treatment, responses differed significantly between groups (P <.001, **E-Figure 7**). Half of patients reported the requirement would have no impact, while 25% said it would make them more likely and 16% less likely to pursue treatment. Among providers, only 27% said the requirement would have no impact; 39% said it would increase and 27% said it would decrease their likelihood of using the treatment. When nutrition visits were framed as optional rather than required, most patients expressed willingness to participate, especially in one-on-one video visits (68% “very likely”). Willingness decreased for group sessions and in-person formats (**E-Figure 8**).

### Sensitivity Analysis

Compared with patients from California, those residing in other locations had similar characteristics (**E-Table 4 & 5**) and shared comparable opinions (**E-Figure 9 & 10**), suggesting that patient responses are broadly representative of patients nationwide.

## DISCUSSION

Our comprehensive survey of patients and sleep medicine providers, primarily from the US, reveals that both CPAP and tirzepatide are broadly acceptable treatment options for COBOSA. Assuming equal effectiveness, patients favored tirzepatide while providers preferred CPAP. Notably, providers with experience prescribing injectable weight-loss medications showed greater alignment with patient preferences, suggesting that clinical familiarity may shift provider views over time. Providers were also more supportive than patients of combination therapy—CPAP plus tirzepatide—if it led to superior outcomes. Given the recent FDA approval of tirzepatide for COBOSA, our findings underscore clinical equipoise and a need for real-world comparative effectiveness data to address a vital question for patients and clinicians alike: should patients with obesity who are newly diagnosed with OSA be initiated on tirzepatide, CPAP or both?^37^

The answer likely depends on patient subgroups. For example, women tend to lose more weight than men with GLP1 receptor agonists;^38^ older adults may experience less weight but more muscle loss;^38–40^ and marginalized or rural populations—who face higher obesity burdens and reduced access to device therapy—may particularly benefit from pharmacotherapy.^3, 41–44^ In the absence of definitive data, treatment decisions are currently shaped by personal preferences and expert opinions.^33^ Our survey adds the voices of key stakeholders to these discussions, informing upcoming clinical guideline updates for the management of COBOSA, and the ongoing design of a pragmatic, patient-centered, comparative effectiveness trial (C<u>PA</u>P and/or Tirzepatide <u>A</u>fter <u>Di</u>a<u>g</u>nosing <u>M</u>ajor <u>Sleep</u> Apnea, PARADIGM-SLEEP trial).

Such a head-to-head trial may shift the “CPAP-first” paradigm to a patient-preferred approach and provide patients access to new therapeutic opportunities—but regardless of the results, it is urgently needed to help optimize care for millions with COBOSA.^37^ SURMOUNT-OSA demonstrated tirzepatide’s efficacy under ideal conditions (including intensive lifestyle counseling), evaluated a limited set of outcomes,^25, 31^ and compared tirzepatide only to placebo—not against CPAP or both.^25^ It remains unclear whether starting both therapies offers an additive benefit or imposes a “double burden”— potentially reducing adherence and straining limited healthcare resources. Furthermore, in routine practice, tirzepatide adherence may fall short of the >80% in the SURMOUNT trials.^24, 25^ In fact, some early real-world studies, including primarily older GLP-1 agents, reported lower adherence,^45, 46^ but these were conducted outside sleep medicine and during periods of limited coverage and shortages.

Notably, greater weight-loss efficacy was associated with improved adherence in one study,^46^ thus supporting optimism for tirzepatide use in practice, consistent with the clinical experiences from some of the authors. Moreover, even with equal reductions in effective AHI, event distributions may differ: tirzepatide may yield a continuous, partial reduction in events, whereas CPAP may provide near-resolution during use but a return to baseline with non-use. These divergent patterns could differentially affect sleep architecture, symptoms, and cardiometabolic outcomes, underscoring the need to assess a broad range of stakeholder-important endpoints beyond OSA severity.

Preserving a healthy fat-to-lean mass loss ratio (about 3:1 in tirzepatide trials^24^) may require diet and exercise support.^47^ However, our results suggest that mandatory in-person nutritionist visits may deter uptake in a substantial proportion of patients and providers. Conversely, other participants—especially many providers—reported a greater likelihood of using tirzepatide if adjunctive nutritional counseling was required. Time constraints may make additional visits particularly challenging for disadvantaged populations (e.g., residents in rural areas, patients with low socio-economic status). A reasonable compromise may be to encourage, but not require routine nutrition visits—preferably as one-on-one sessions with video options, which were highly acceptable to patients. Additionally, freely available educational videos tailored to tirzepatide use in COBOSA may further support scalable and equitable access to medication and lifestyle counselling. In combination with dedicated training and evidence-based protocols, such educational materials may also empower sleep clinicians with limited experience prescribing injectable obesity medications, who otherwise appear hesitant to use tirzepatide. Such resources could be developed as part of the envisioned PARADIGM-SLEEP trial and help with the rapid translation of optimal, personalized treatment strategies into practice.

Historically, OSA management has followed a one-size-fits-all approach, with CPAP as the default therapy. However, there is growing recognition of clinically meaningful subgroups based on pathophysiology (endotypes) and consequences (phenotypes).^48^ For instance, patients with comorbid insomnia and sleep apnea (COMISA) benefit from tailored treatment strategies.^49, 50^ We believe COBOSA constitutes a similarly distinct subgroup with a high prevalence—estimated to include 40– 60% of OSA patients. Our provider data support estimates on the higher end of that range, reinforcing the importance of this subtype. Like COMISA, COBOSA likely warrants individualized treatment and focused research to improve outcomes.

### Strengths and Limitations

This study engaged two key stakeholder groups—patients and sleep medicine providers—during a time of active discourse on how to best incorporate weight-loss pharmacotherapy into the care of patients with COBOSA. The provider sample was geographically and demographically diverse, with representation from various practice settings. While most patient responses came from California, sensitivity analyses showed that non-California participants had comparable characteristics and attitudes, supporting the generalizability of findings. Of note, California responses also tended to be collected earlier in the study, while responses from other states came later. The consistency between these two groups further suggests that patient attitudes remained stable over time.

A major limitation is the potential for selection bias. For instance, of the 6,626 email invitations to participate in the survey, about 5% responded, which may limit generalizeability of our findings to those having easy web-access and interest to share their views. Future work should consider multimodal outreach efforts to maximize response rates,^51^ particularly among populations that have been historically suboptimally treated for COBOSA despite being disproportionately more affected, such as those living in rural areas.

Furthermore, as with any open online survey, duplicate or bot responses are a theoretical risk. However, use of the Google reCAPTCHA feature, quality control measures (e.g., survey duration times, hidden fields), the absence of financial incentives, and the timing of responses linked closely to specific outreach efforts collectively support high data integrity.

Finally, providers with more experience prescribing injectable weight-loss medications were more likely to favor tirzepatide, but given the cross-sectional study design it is unclear whether clinical experience led to greater openness—or whether providers already inclined to favor tirzepatide were more likely to gain that experience (i.e., possible reverse causation). Future longitudinal studies should assess whether targeted provider training influences treatment attitudes and prescribing behavior.

## Conclusion

COBOSA represents a large and clinically significant subgroup of patients with OSA. Both tirzepatide and/or CPAP appear to be broadly acceptable treatment options for patients and providers. However, real-world comparative effectiveness data are needed to determine the optimal treatment strategy for the millions of individuals affected by COBOSA.

## Data Availability

The dataset analyzed in this study is de-identified by design. Access can be granted upon reasonable request to the corresponding author, contingent on approval by the UCSD IRB and completion of a data use agreement.

## DECLARATIONS

### Ethics approval and consent to participate

The UCSD IRB determined that this study (#811561) met the criteria for exempt research under 45 CFR 46.104(d) 2 (i), and granted a partial waiver to use protected health information to email invitations to select UCSD sleep clinic patients. The landing page of the online survey outlined the survey’s purpose, eligibility criteria, estimated completion time, types of information queried, and emphasized that participation was voluntary and anonymous, with completion implying consent.

## Funding

Dr Schmickl is supported by the National Institutes of Health (NIH; K23HL161336). Dr Kundel is supported by the NIH K23HL161324, and the American Academy of Sleep Medicine Foundation (AASMF 274-BS-22). Dr Zinchuk is supported by the NIH (K23HL159259, R01HL179077). Dr Nokes is supported by the Department of Veterans Affairs CSR&D CDA-2 IK2CX002524-01A2 and NIH Loan Repayment Program. Dr Orr is supported by the NIH (K23 HL151880). Dr Martin is supported by a VA Senior Research Career Scientist Award (HSR MRA0-004-24W), and NIH/NHLBI K24 HL143055. Dr Malhotra is funded by NIH.

The project described was partially supported by the NIH, Grant UL1TR000100 of CTSA funding prior to August 13, 2015 and Grant UL1TR001442 of CTSA funding beginning August 13, 2015 and beyond. The content is solely the responsibility of the authors and does not necessarily represent the official views of the NIH.

### Authors’ contributions

All authors contributed substantially to the study conception and design (CNS, AT, JM, AM), data acquisition (CNS, AT, BM, MM, BN, VK, MT, RSA, AZ, TZ, MSB, RLO, AM), analysis (CNS), and/or interpretation (all) of this study. CNS drafted the manuscript, and all authors revised it critically for intellectual content. All authors gave final approval of this version to be submitted.

## Acknowledgements

We would like to thank our PARADIGM-SLEEP collaborators: Dr Michelle Zeidler and Ms Kimberly Fox (University of California, Los Angeles, CA); Dr Ken He and Mr Martin O’Donnell (VA Puget Sound, Seattle, WA); Dr Neomi Shah and Mr Luis Dejesus (Mt Sinai New York, NY); Dr Klar Yaggi, Dr Brian Wojeck, Dr Ania Jastreboff and William Wivel (Yale University, New Haven, CT); Dr Julie Neborak, Dr Lindsay McCullough, and Ms Nancy Vitucci (Rush University, Chicago, OH); Dr Venkatesh Krishnamurthy, Ms Emma Oldham, and Ms Debbie Hlasnik (University of Pittsburgh, PA); Dr Kara Dupuy-McCauley, Dr Melissa Lipford and Mr Jon Morphew (Mayo Clinic Rochester, MN), Dr Richard Bogan, Dr Laura Herpel, and Ms Jane Lewis (Bogan Sleep Consultants, Columbia, SC), Dr Abdulghani Sankari, Dr Salam Zeineddine, and Mr Dion Williams (Wayne State University, Detroit, OH); Dr Reena Mehra, Dr Martha Billings, Dr Sina Gharib, and Dr Vishesh Kapur (University of Washington, Seattle, WA), Ms Ginny Meyerhuber (VA San Diego, CA); Dr Yulia Lokhnygina (Duke University School of Medicine, Durham, NC); Dr Marc Raphaelson (VA West Virginia); Dr Dayna Johnson (Emory University); Dr Biren Kamdar, Dr Karen McCowen, Ms Pam Tilden, Ms Pamela DeYoung, and Ms Stacie Moore (University of California, San Diego); Dr Ali Azarbarzin (Harvard University, Boston, MA); Dr Linda Gallo (San Diego State University, San Diego, CA).

## ONLINE SUPPLEMENT

**E-Figure 1.**
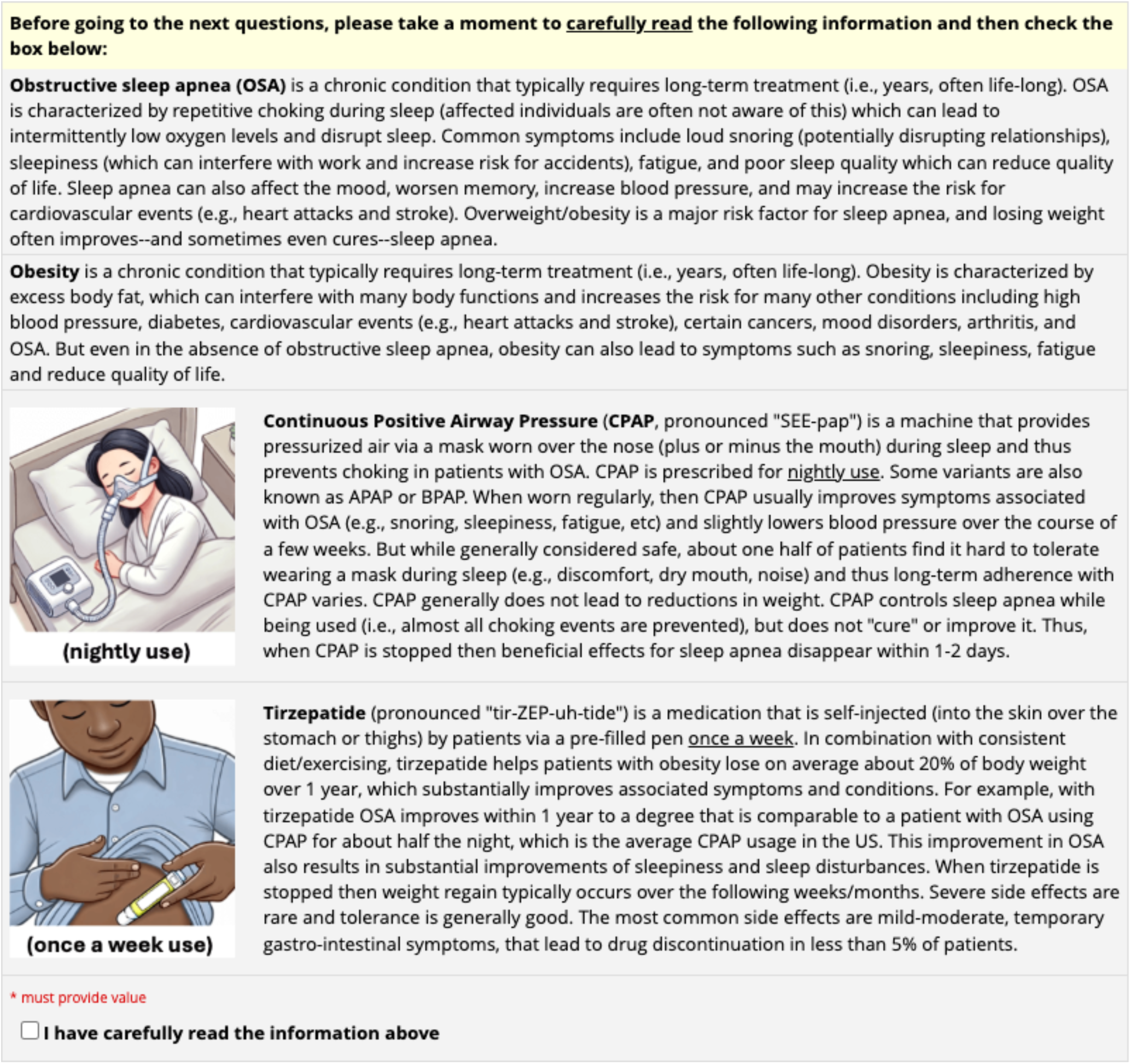
Disease and Treatment Overview. Provided to Participants prior to questions about acceptability/preferences for CPAP and/or Tirzepatide.

**E-Figure 2.**
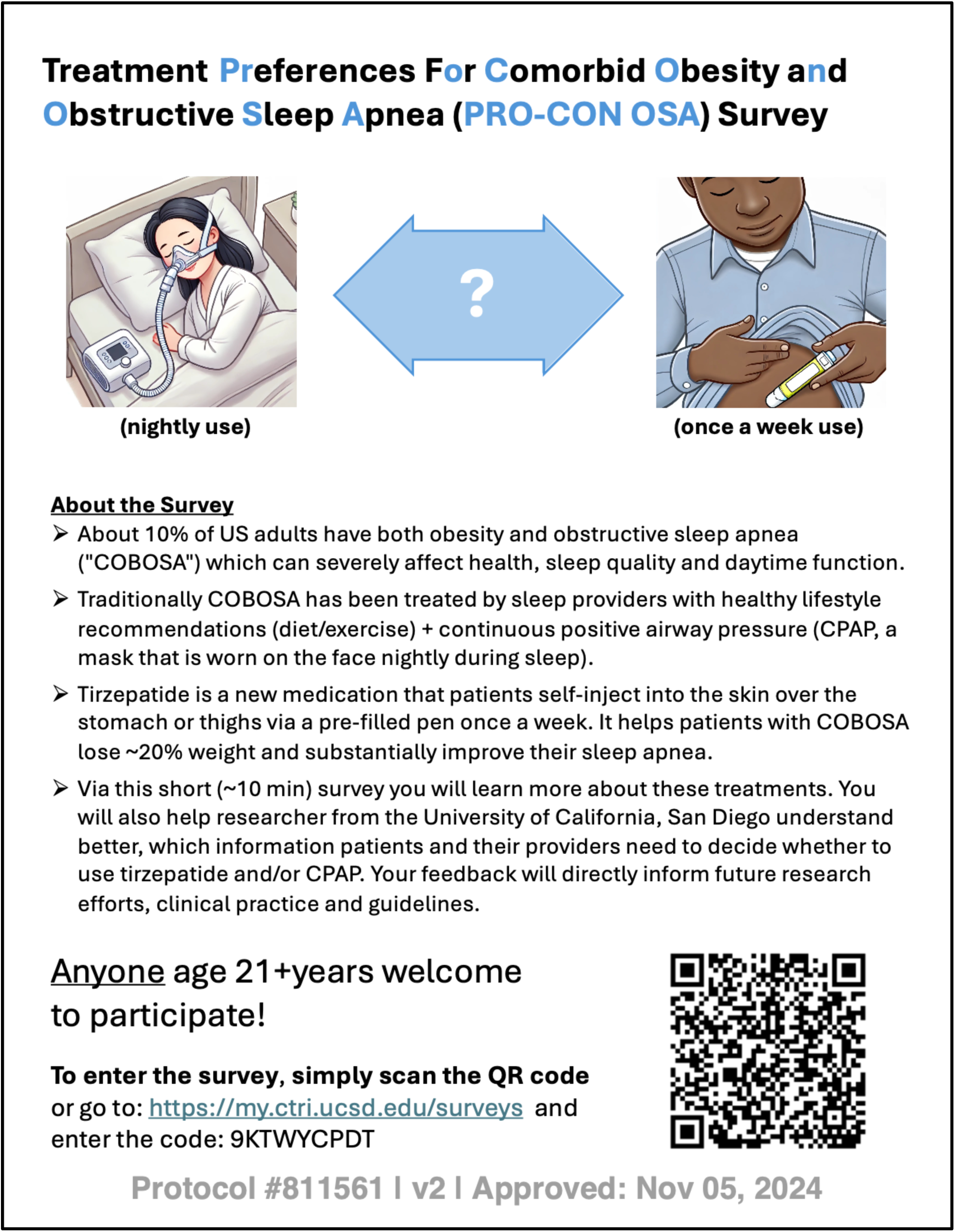
Flyer A.

**E-Figure 3.**
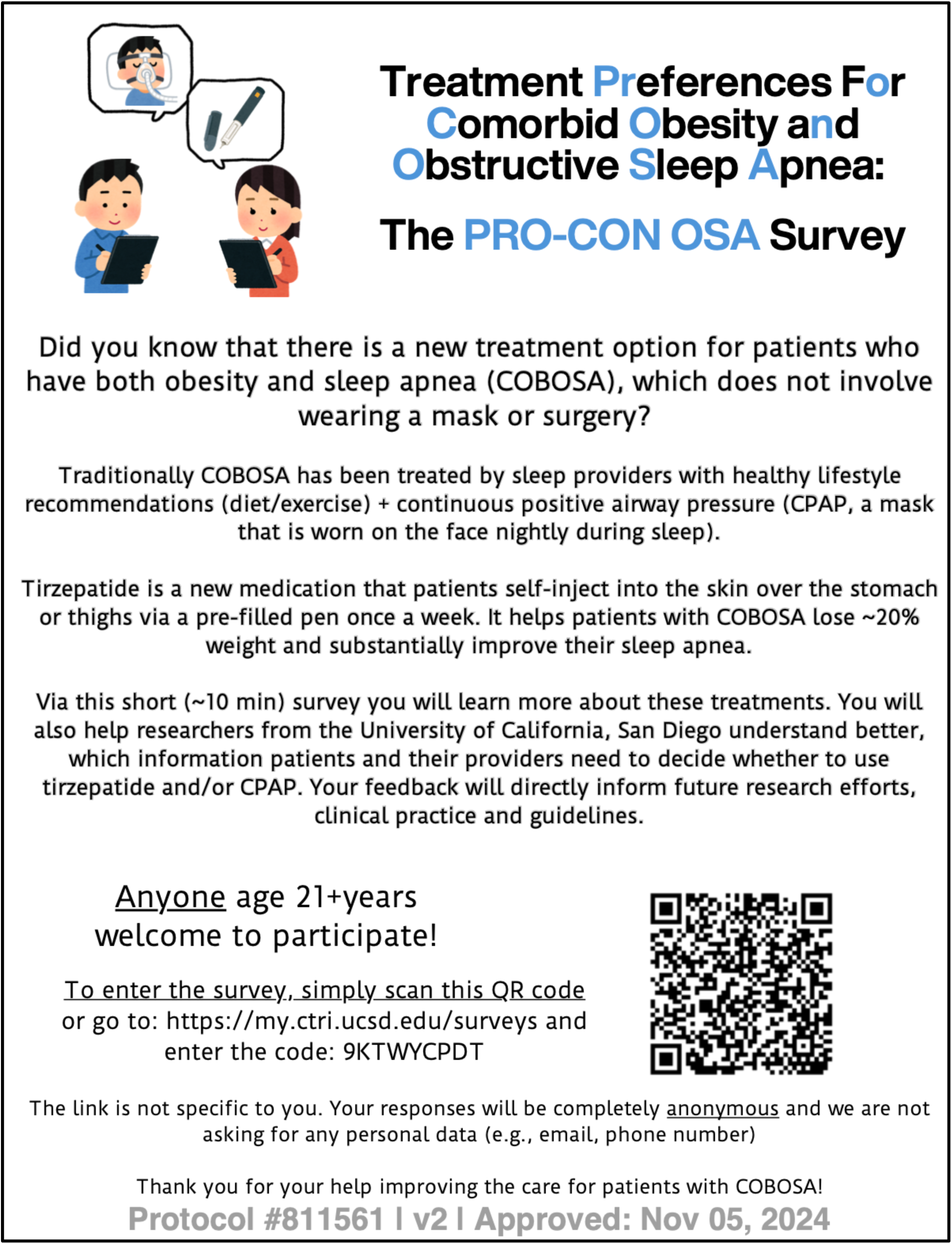
Flyer B.

**E-Figure 4.**
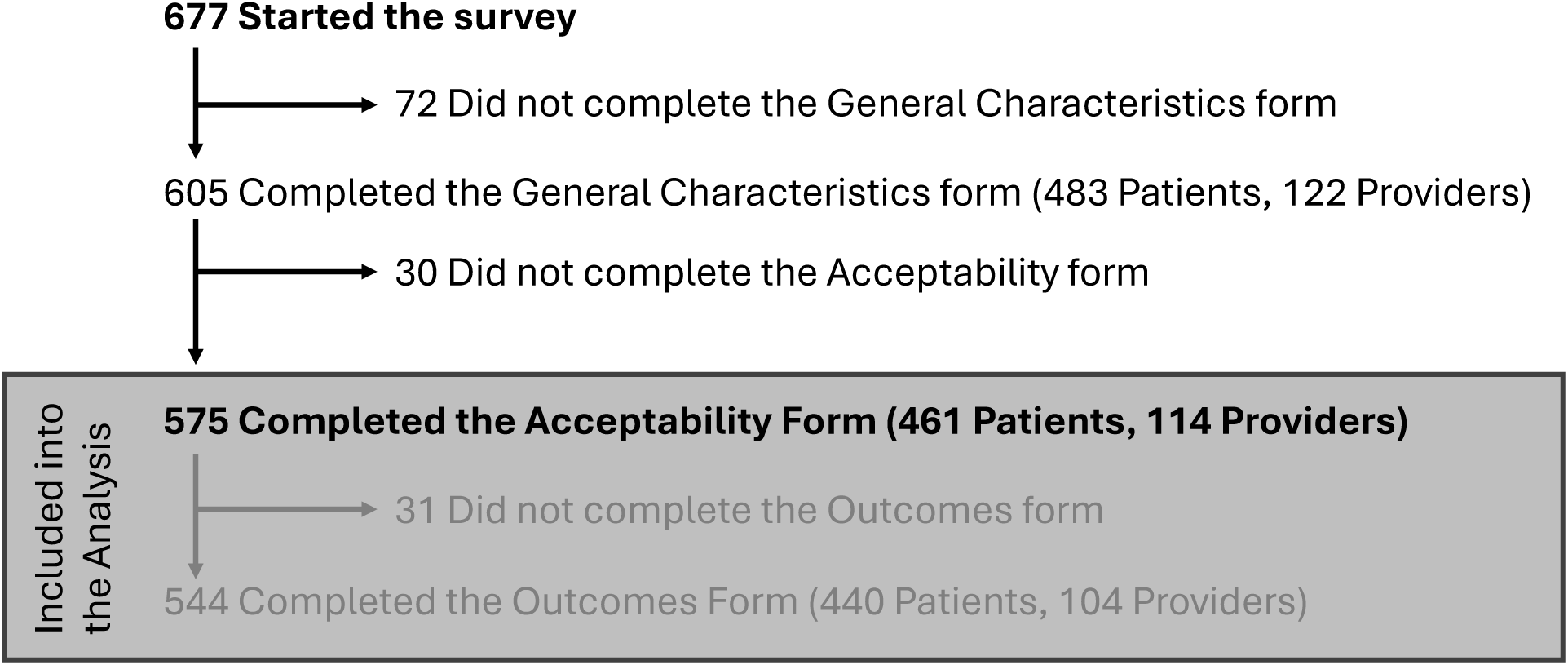
Flowchart.

**E-Figure 5.**
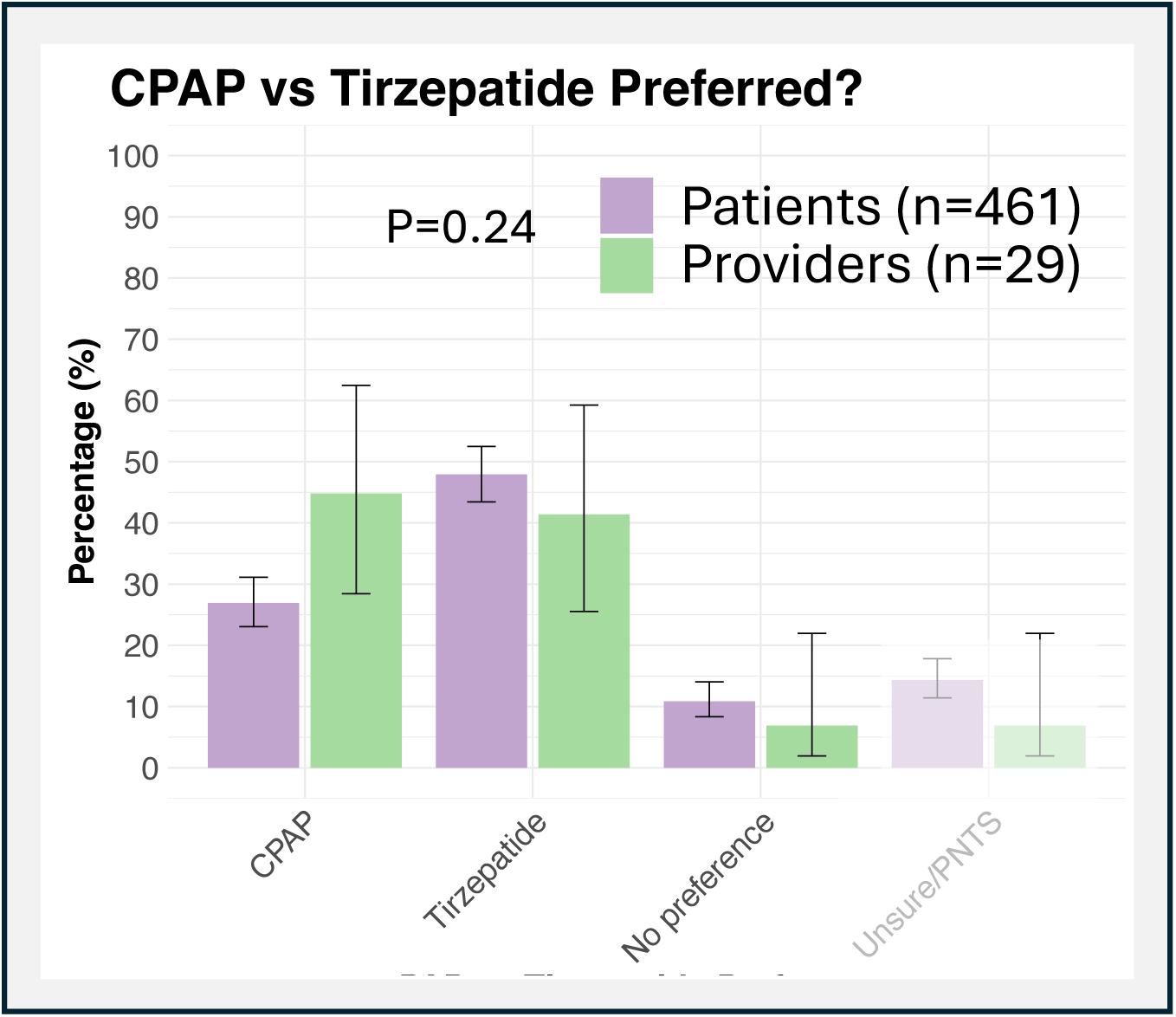
CPAP vs Tirzepatide Preference: Patients vs 29 Providers <u>with</u> Substantial Experience Using Injectable Weight Loss Medications. If there was strong evidence that both Tirzepatide and CPAP treat sleep apnea and associated risks/ symptoms similarly well, which one would you prefer long-term? Provider responses are green, non-provider (i.e., patients) responses are purple. Response patterns did not significantly differ between patients and providers across all four assessments (Fisher’s exact test, P = 0.24). Bars reflect 95% confidence intervals. PNTS = Prefer not to say.

**E-Figure 6.**
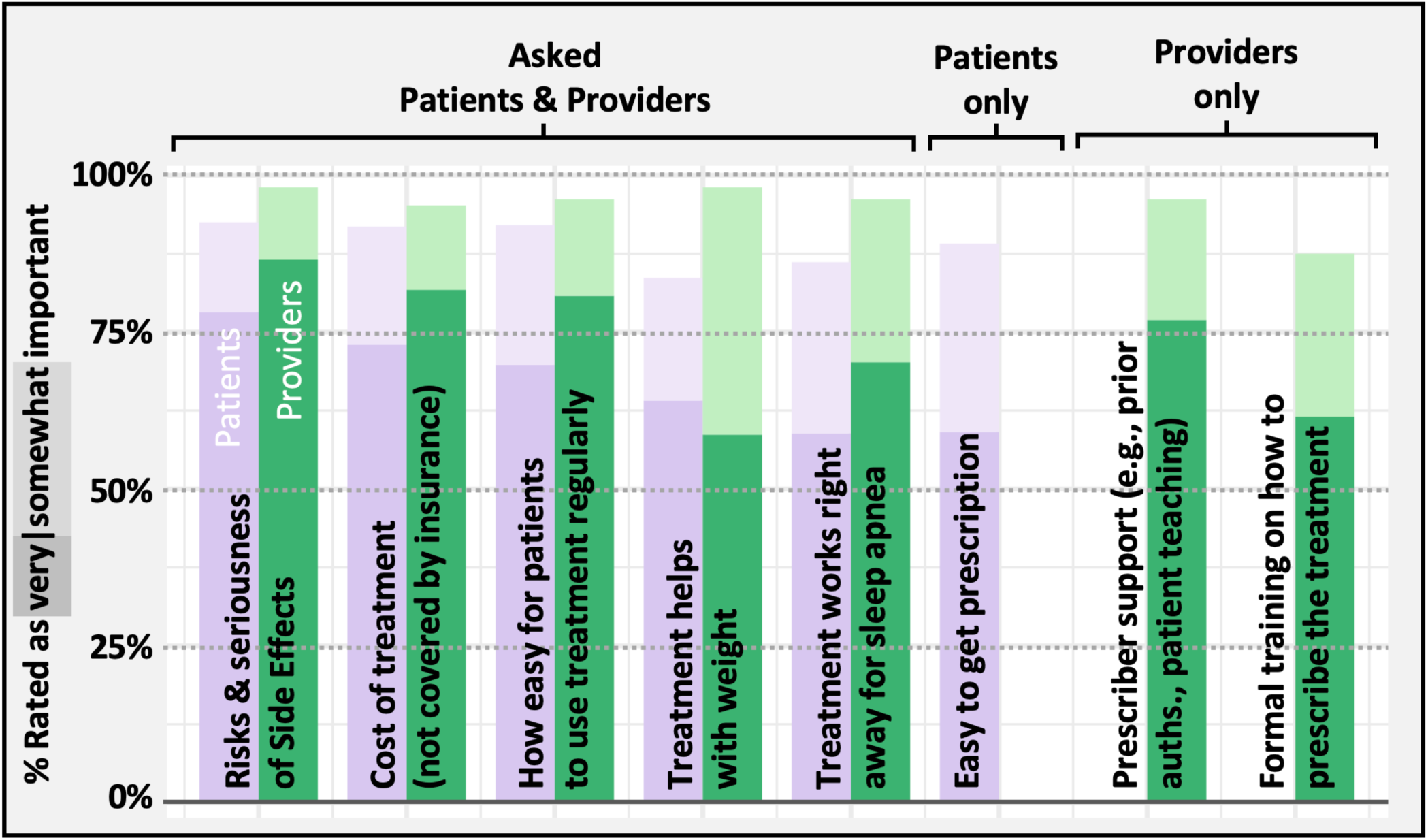
Other Factors Important for Decision Making. If you had (a patient with) newly diagnosed COBOSA: When trying to decide whether to use Tirzepatide and/or CPAP for OSA, how important would be the following factors for your decision? Provider responses are green, non-provider (i.e., patients) responses are purple. “Very” important = dark shade, “Somewhat” important = light shade.

**E-Figure 7.**
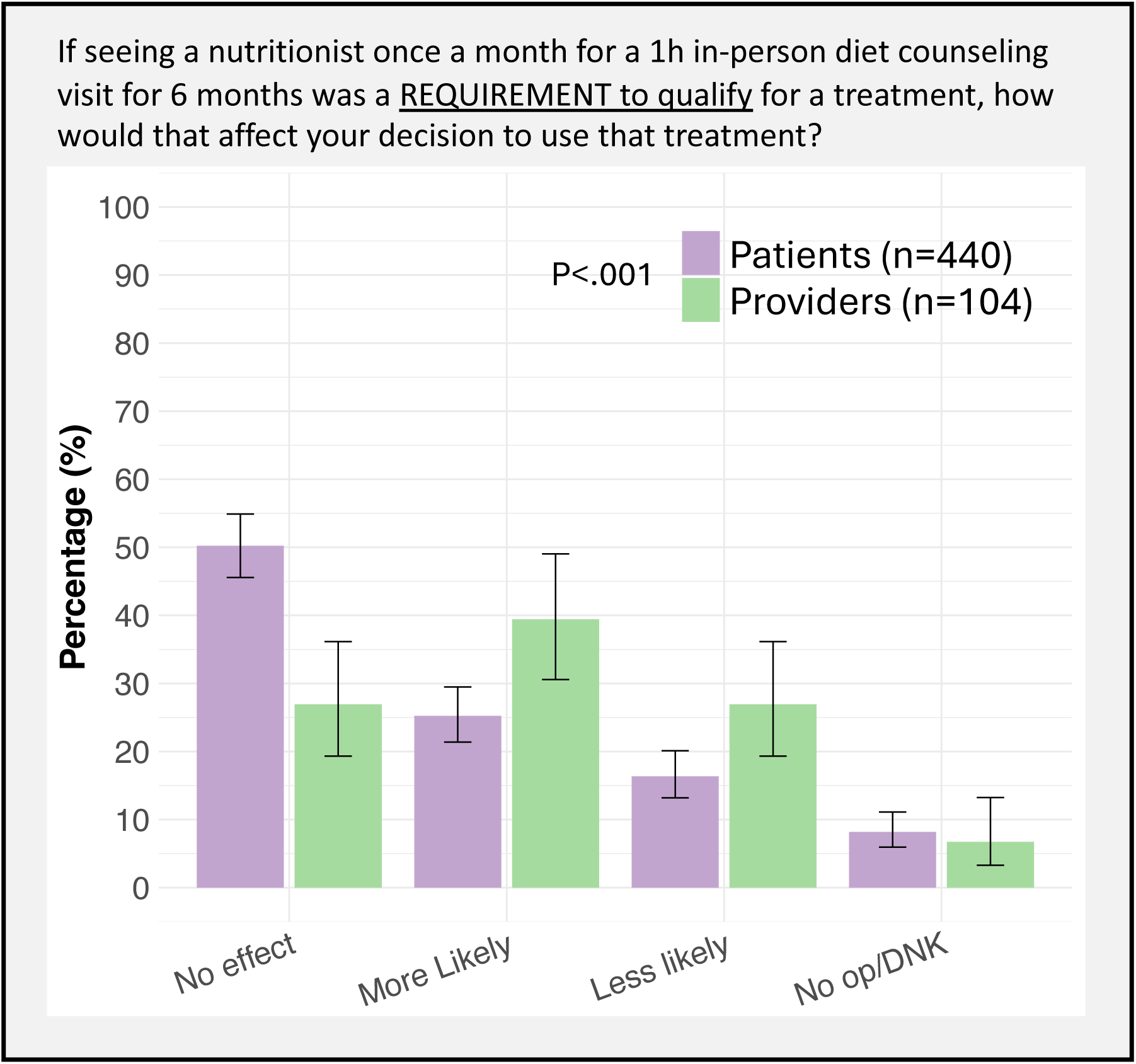
Impact of Nutritionist Requirement on Therapy Use: Patients vs Providers.

**E-Figure 8.**
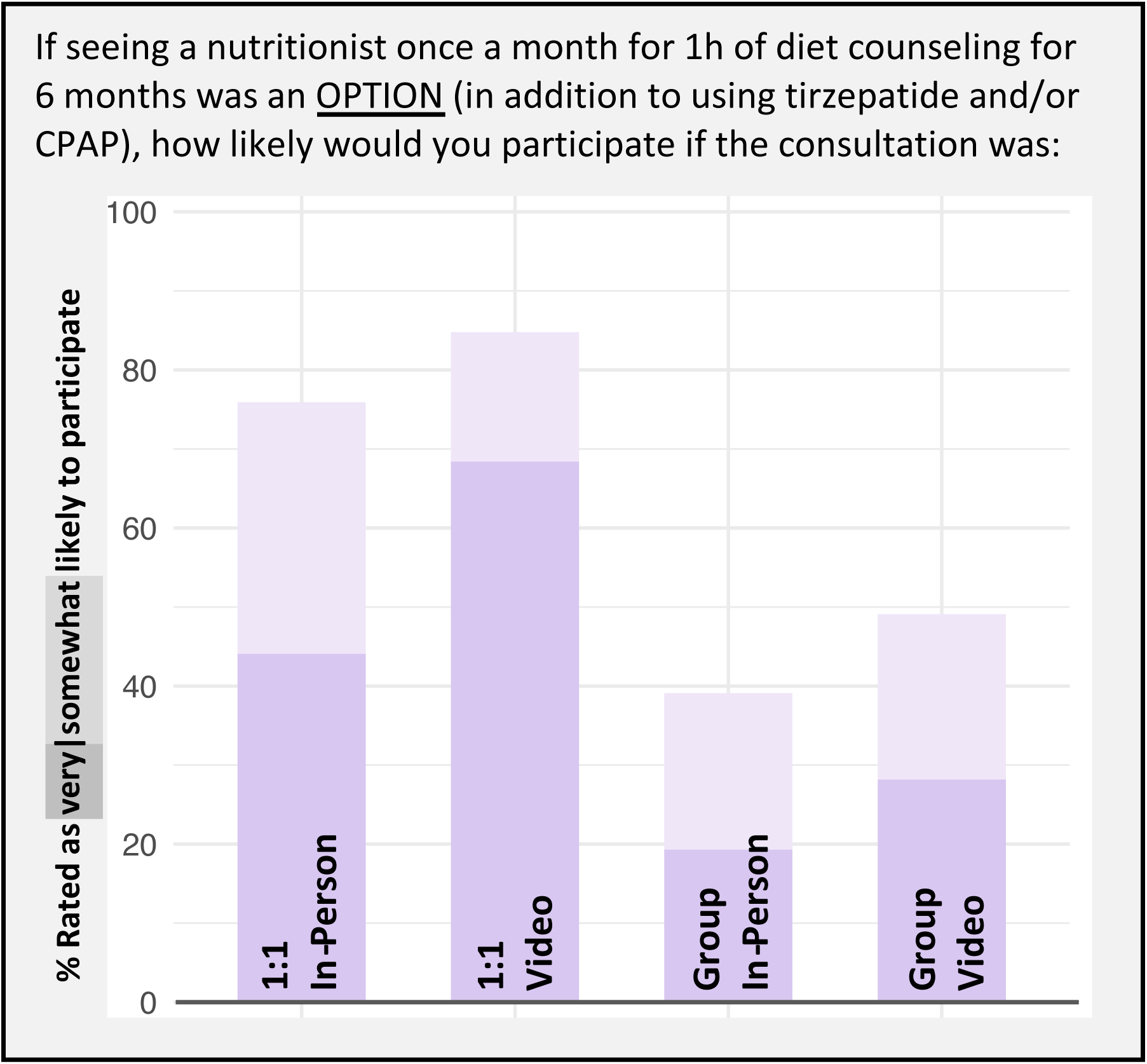
Patient Preferences for Optional Nutrition Visit Settings.

**E-Figure 9.**
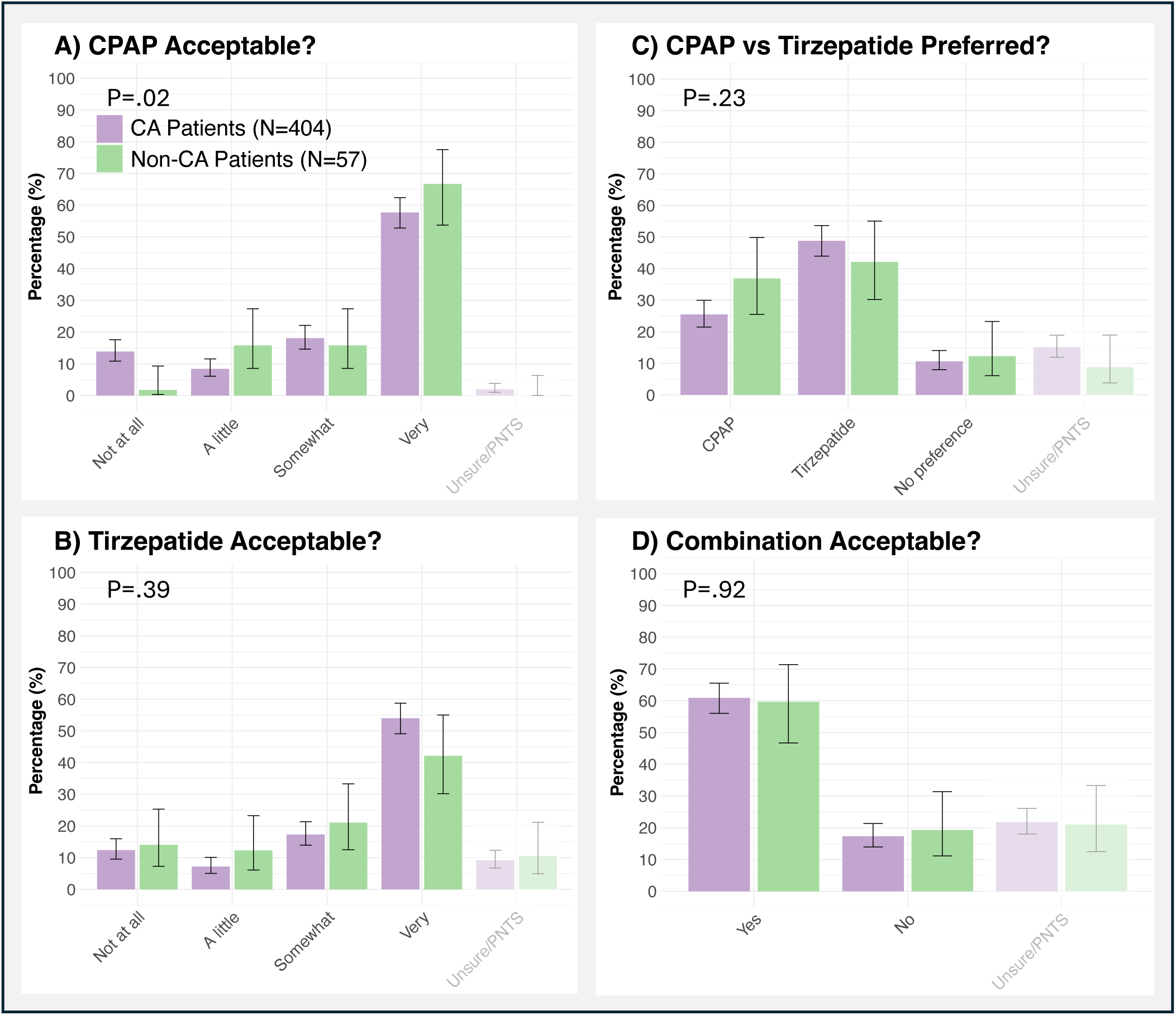
Sensitivity Analysis: Acceptability and Preferences Patients from CA vs non-CA. If you had (a patient with) newly diagnosed, untreated COBOSA, how acceptable would you find it to use/prescribe CPAP (**Panel A)** or tirzepatide **(Panel B**). If there was strong evidence that both Tirzepatide and CPAP treat sleep apnea and associated risks/ symptoms similarly well, which one would you prefer long-term? (**Panel C**). If there was strong evidence that combining both CPAP + Tirzepatide leads to greater improvements of sleep apnea and associated risks/symptoms than using either CPAP or tirzepatide alone, would you be willing to use/prescribe both of them together long-term? (**Panel D**). Responses from patients residing in California (CA) are purple, while responses from patients living outside of California (i.e., Non-CA patients) are green. Response patterns did not differ significantly between CA and non-CA patients except for CPAP acceptability (Fisher’s exact test). Bars reflect 95% confidence intervals. PNTS = Prefer not to say.

**E-Figure 10.**
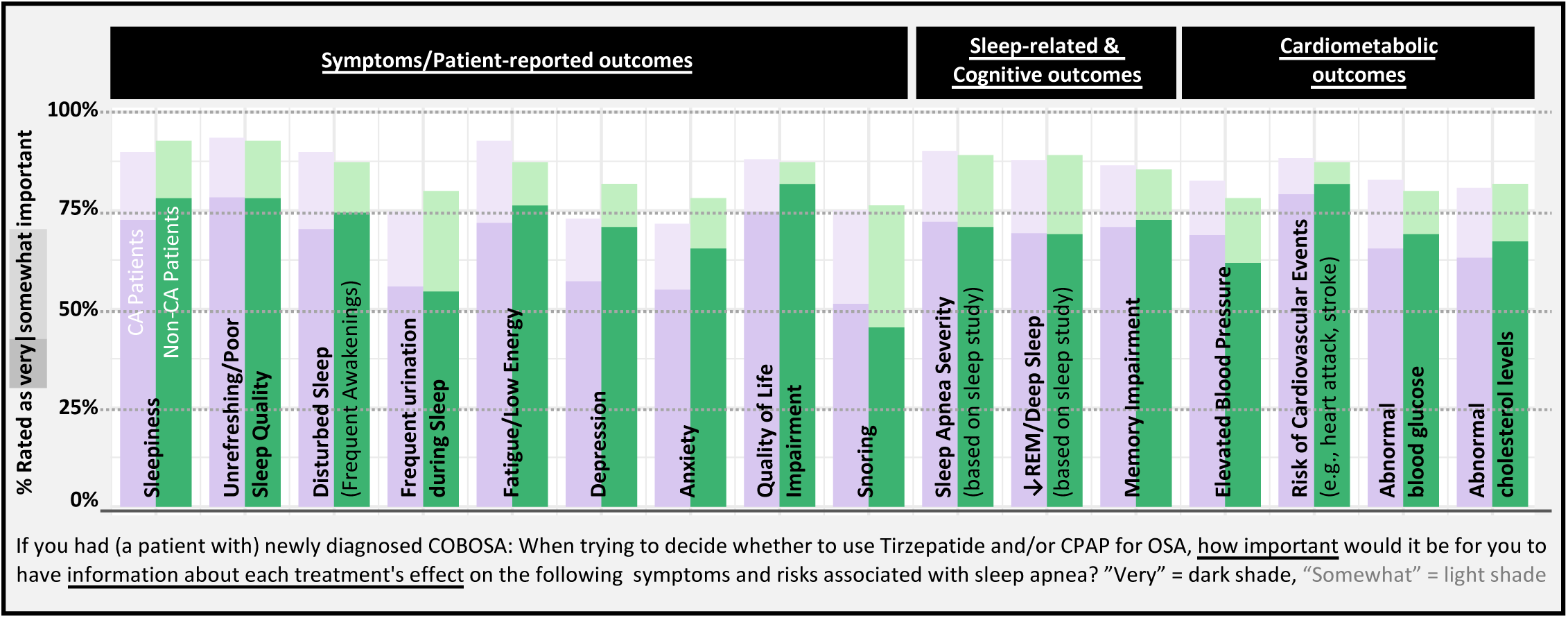
Sensitivity Analysis: Importance of Information for Patients from CA vs non-CA. Responses from patients residing in California (CA) are purple, while responses from patients living outside of California (i.e., Non-CA patients) are green. Ratings were overall similar. “Very” important = dark shade, “Somewhat” important = light shade.

**E-Table 1.** Tirzepatide’s Effect on OSA vs CPAP Alone. Most outcomes data on tirzepatide in the table below are from the original SURMOUNT-OSA publication (Malhotra et al., NEJM 2024)^1^ which assessed “key secondary outcomes” at 1 year follow up, and tested them sequentially to preserve type 1 error. Subsequently, an article (Kanu et al., Sleep Med 2025)^2^ on other secondary/exploratory outcomes (i.e., no adjustment for multiple testing) reported statistically significant improvements with tirzepatide vs placebo in some domains (activity, vigilance) of the Functional Outcomes of Sleep Questionnaire (FOSQ), in all but the social functioning domain and mental component summary scores of the Short-Form 36 Health Survey, Version 2 (SF-36v2), in the health status score and visual analog scale of the EQ-5D-5 Level (EQ-5D-5L), in global impression of severity/change scales for fatigue, sleepiness, sleep quality and snoring. Generally, changes in FOSQ, EQ-5D-5L and SF-36 following tirzepatide were comparable to changes expected with CPAP use.^3–6^ Epworth sleepiness score (ESS) improved by a statistically significant amount in trial 1 (no concomitant CPAP; –1.4 95%-CI –2.5 to –0.3) but not in trial 2 (concomitant CPAP since baseline; –0.9, 95%-CI –2.1 to –0.2), with changes smaller than typically seen with CPAP.^7^

|  | **SURMOUNT-OSA<sup>1, 2</sup>**<br>(Malhotra et al., NEJM 2024; Kanu et al. Sleep Med 2025) |  | **Typical Effects**<br>**Observed With**<br>**CPAP Alone (vs control)** |
| --- | --- | --- | --- |
|  | **Tirzepatide<sup>a</sup>** | **Placebo<sup>a</sup>** |  |
| Weight (%) | -18 to -20%<sup>a</sup> | -2% | Not much change<sup>8, 9</sup> |
| Effective AHI (%)<sup>b</sup> | -51 to -59%<sup>a</sup> | -3% | -40 to -60%<sup>d 10</sup> |
| ESS (raw score)<sup>c</sup> | -3.3 to -3.6 | -1.9 to -2.6 | -2 to -3<sup>7</sup> |
| PROMIS-SRI (T-score)<sup>c</sup> | -6.6 to -8.2<sup>a</sup> | -3.1 to -3.9 | -4.2<sup>11 e</sup> |
| PROMIS-SDA (T-score)<sup>c</sup> | -4.5 to -7<sup>a</sup> | -2.4 to -3.1 | -4.0<sup>11 e</sup> |
| FOSQ (raw score) | +1.5 to 2.0 | +0.9 to 1.7 | +1<sup>5</sup> |
|  |  |  | (+0.3 effect sizes in meta-analyses)<sup>7</sup> |
| Systolic BP (mmHg)<sup>c</sup> | -7.6 to -9.5<sup>a</sup> | -1.8 to -3.9 | -2 to -3<sup>7, 12</sup> |
| Diastolic BP (mmHg) | -3.3 to -4.9<sup>a</sup> | -2.1 to -2.2 | -1 to -2<sup>7, 12</sup> |
| CRP<sup>c</sup> | -1.4 mg/L<sup>a</sup> | -0.3 to -0.7 mg/L | 0 to -20%<sup>13, 14 f</sup> |
|  | (approx. -40%) | (approx. -10 to -20%) | (-0.5 effect sizes in meta-analyses)<sup>15, 16</sup> |

<sup>a</sup> Ranges are from two trials in patients with OSA who were untreated (trial 1) or already on CPAP prior to randomization (trial 2; in this trial CPAP was withheld 7 days prior to any outcomes assessment, thus measurements reflect primarily the tirzepatide effect); comparisons vs placebo were statistically significant for all listed outcomes except ESS in trial 2 and FOSQ;
<sup>b</sup> Primary outcome in SURMOUNT-OSA
<sup>c</sup> “Key secondary outcomes” in SURMOUNT-OSA at 1 year follow up, tested sequentially to preserve type 1 error
<sup>d</sup> Effective Apnea-Hypopnea-Index (AHI) accounting for CPAP adherence is defined as $AHI_{OnCPAP} \times \%TST_{OnCPAP} + AHI_{OffCPAP} \times \%TST_{OffCPAP}$ .<sup>17</sup> The range is based on studies reporting that CPAP reduces the AHI by 80-90% while worn (i.e., reduction in $AHI_{OnCPAP}$ ),<sup>7</sup> and that average CPAP use is typically 3-5h/night,<sup>18-20</sup> corresponding to approximately 40 to 70% of the total sleep time ( $\%TST_{OnCPAP}$ ). Note, for tirzepatide $\%TST_{OnTirzepatide} = 100\%$ and thus the AHI observed during a sleep study following tirzepatide therapy represents the effective AHI.
<sup>e</sup> Study of patients with diabetes screened for OSA; thus, effect estimates for CPAP are likely an underestimate of its effect in typical sleep clinic populations
<sup>f</sup> Another large and potentially informative study by Chirinos et al (NEJM 2014) lacked a control group but found no significant change in CRP from baseline following 24 weeks of CPAP<sup>8</sup>
Abbreviations: SRI = Sleep related impairment, SDA = Sleep disturbance, BP = Blood pressure, CRP = C-reactive Protein (marker of cardiovascular risk). For additional abbreviations see Table legend.

**E-Table 2.**
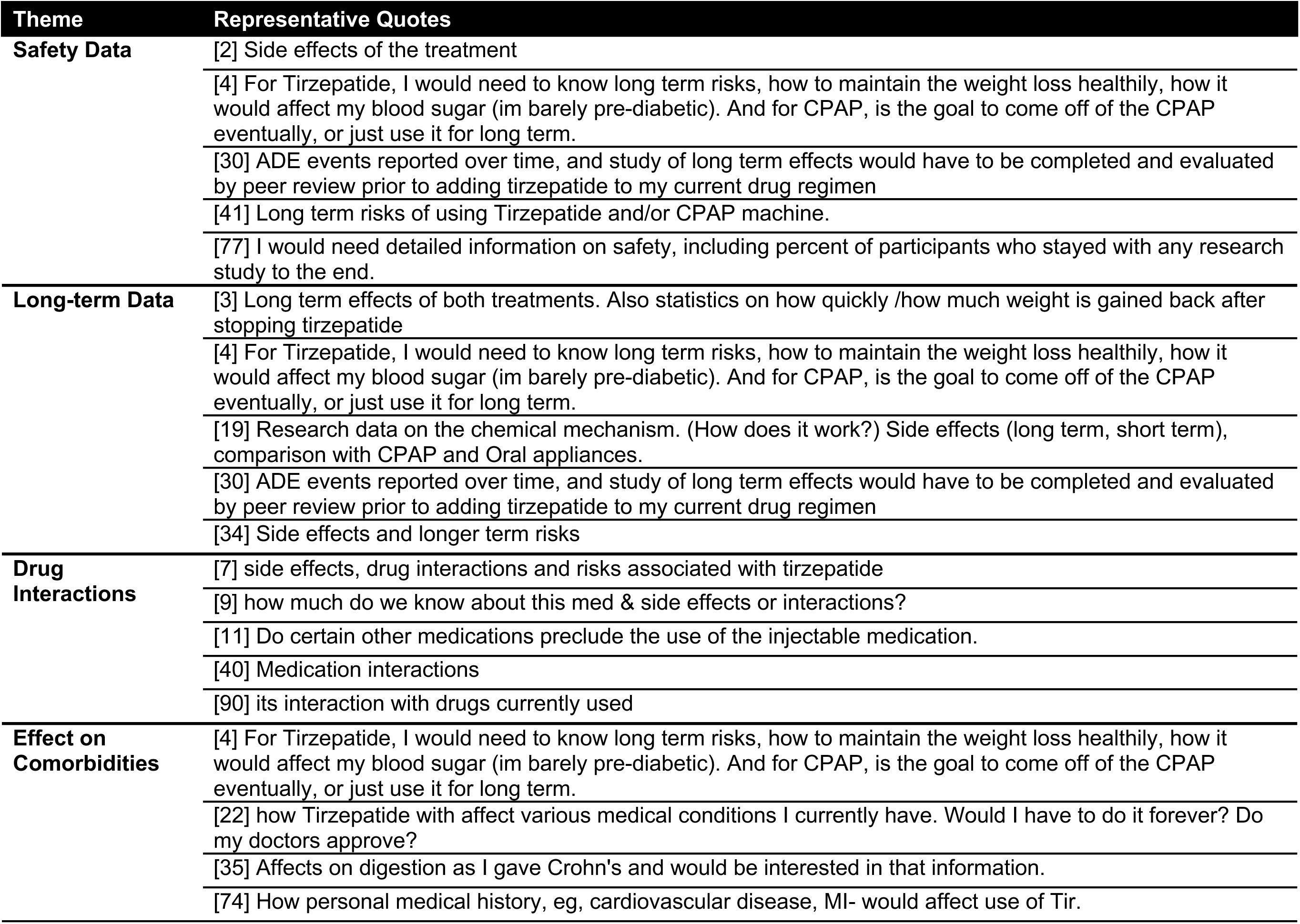
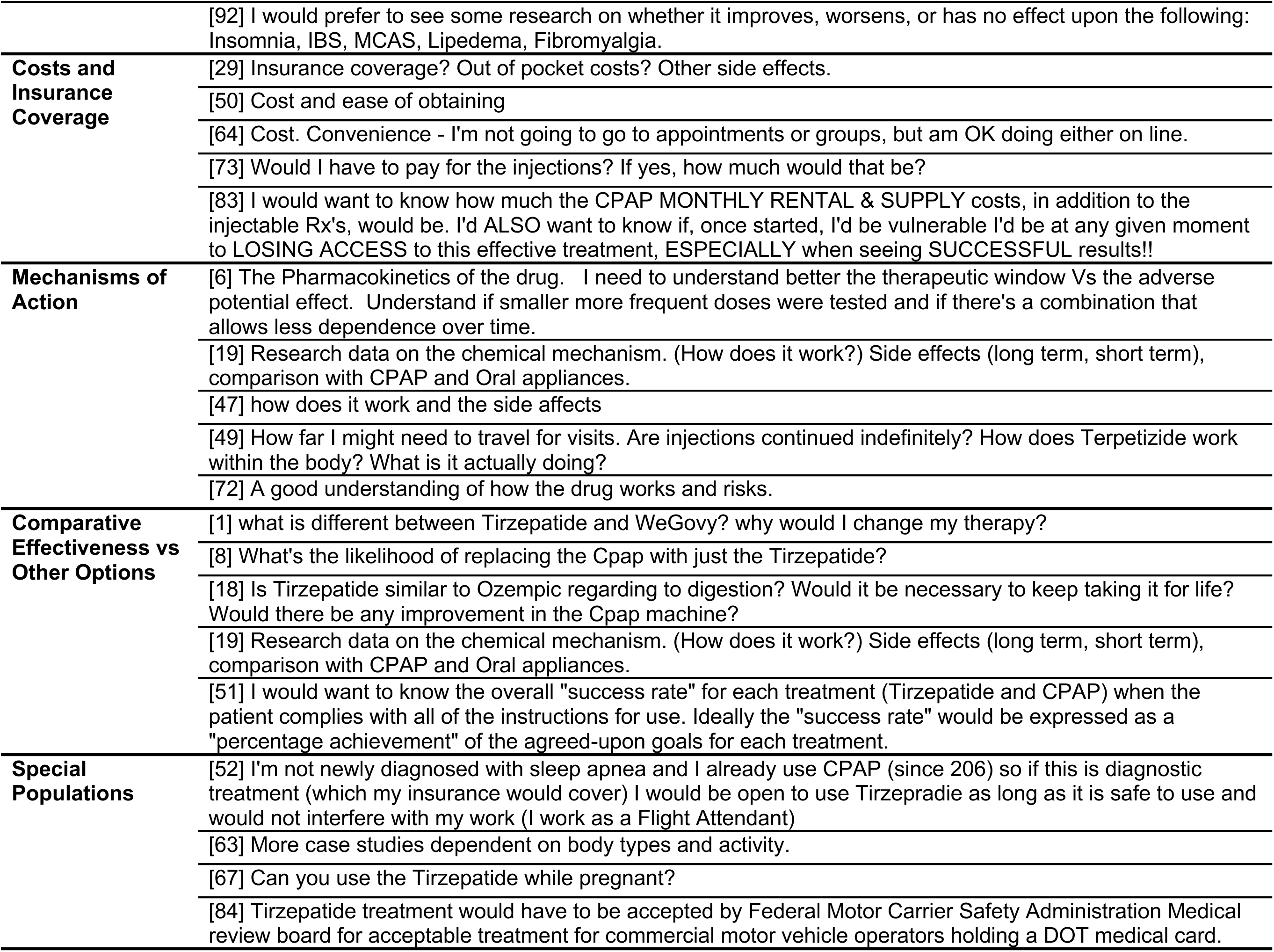
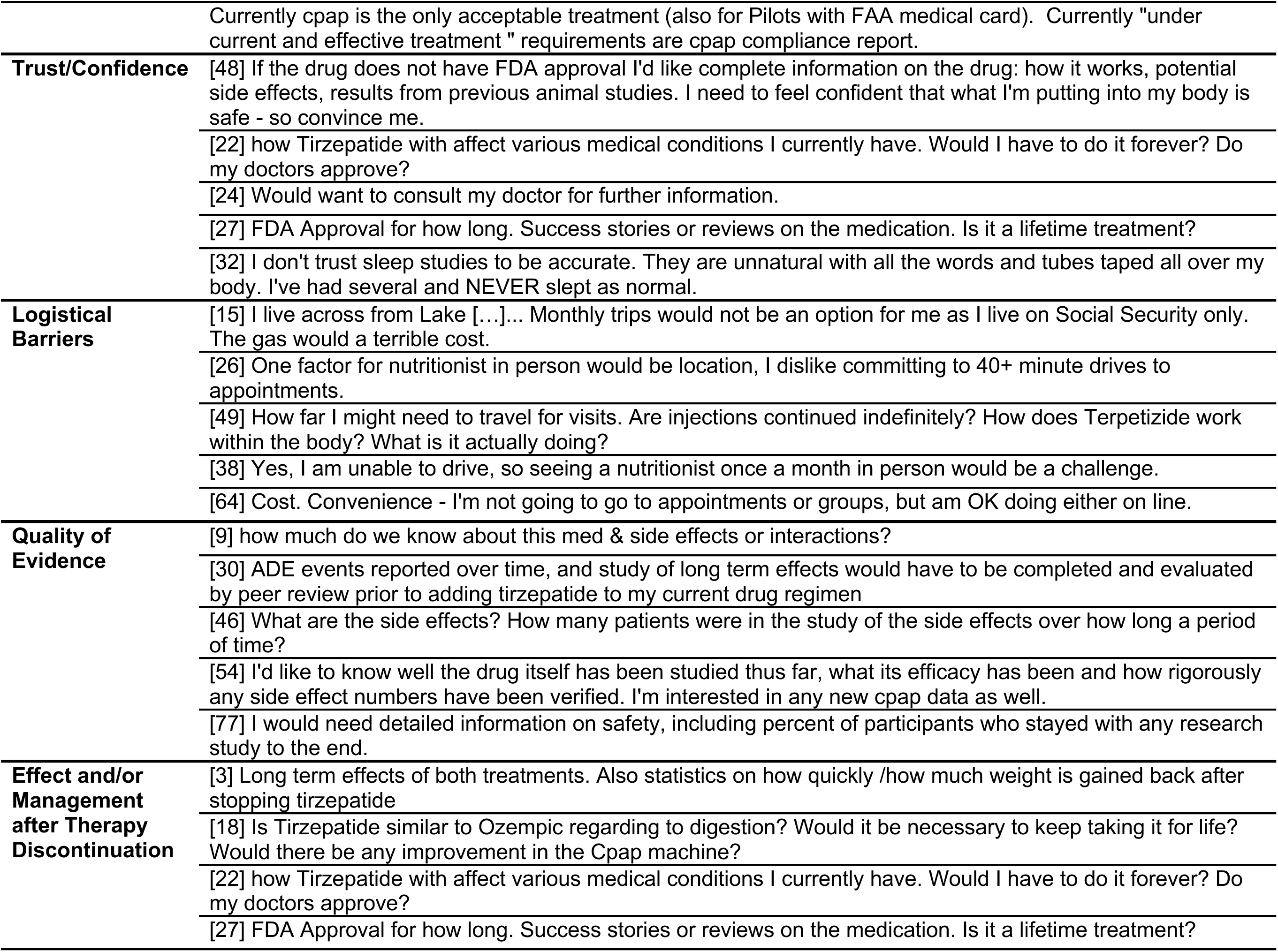
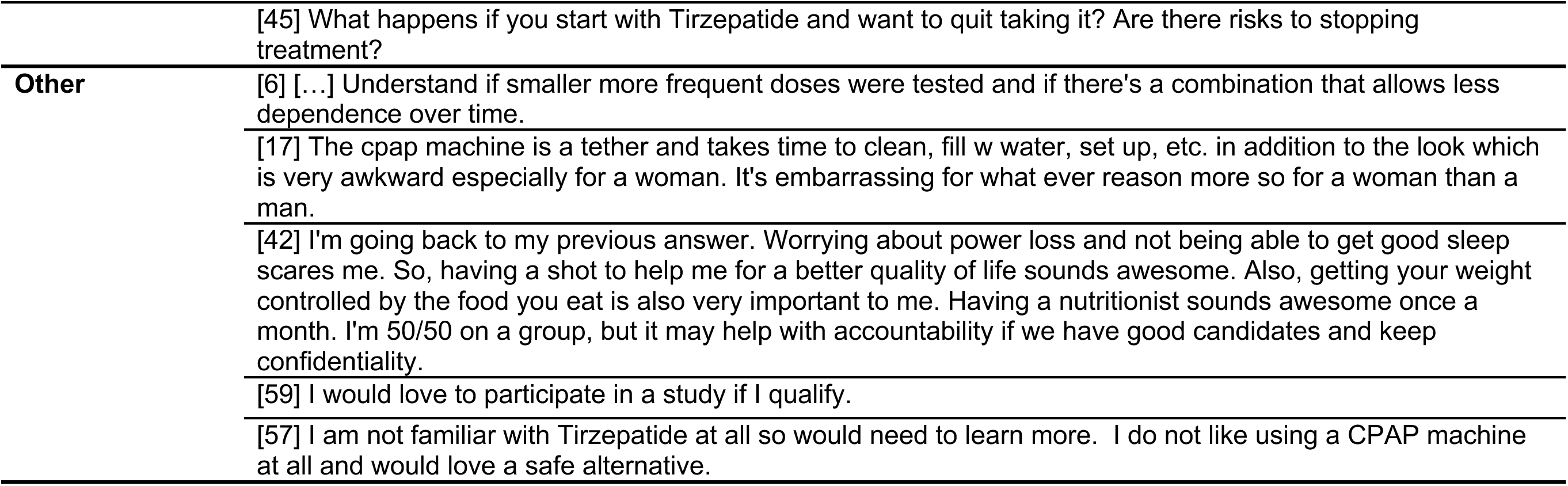
Other Information Needs by Patients: Themes and Representative Quotes. 92 Patients provided free-text comments, some of which related to more than one theme (i.e., 132 related themes).

| Theme | Representative Quotes |
| --- | --- |
| **Safety Data** | [2] Side effects of the treatment |
|  | [4] For Tirzepatide, I would need to know long term risks, how to maintain the weight loss healthily, how it would affect my blood sugar (im barely pre-diabetic). And for CPAP, is the goal to come off of the CPAP eventually, or just use it for long term. |
|  | [30] ADE events reported over time, and study of long term effects would have to be completed and evaluated by peer review prior to adding tirzepatide to my current drug regimen |
|  | [41] Long term risks of using Tirzepatide and/or CPAP machine. |
|  | [77] I would need detailed information on safety, including percent of participants who stayed with any research study to the end. |
| **Long-term Data** | [3] Long term effects of both treatments. Also statistics on how quickly /how much weight is gained back after stopping tirzepatide |
|  | [4] For Tirzepatide, I would need to know long term risks, how to maintain the weight loss healthily, how it would affect my blood sugar (im barely pre-diabetic). And for CPAP, is the goal to come off of the CPAP eventually, or just use it for long term. |
|  | [19] Research data on the chemical mechanism. (How does it work?) Side effects (long term, short term), comparison with CPAP and Oral appliances. |
|  | [30] ADE events reported over time, and study of long term effects would have to be completed and evaluated by peer review prior to adding tirzepatide to my current drug regimen |
|  | [34] Side effects and longer term risks |
| **Drug Interactions** | [7] side effects, drug interactions and risks associated with tirzepatide |
|  | [9] how much do we know about this med & side effects or interactions? |
|  | [11] Do certain other medications preclude the use of the injectable medication. |
|  | [40] Medication interactions |
|  | [90] its interaction with drugs currently used |
| **Effect on Comorbidities** | [4] For Tirzepatide, I would need to know long term risks, how to maintain the weight loss healthily, how it would affect my blood sugar (im barely pre-diabetic). And for CPAP, is the goal to come off of the CPAP eventually, or just use it for long term. |
|  | [22] how Tirzepatide with affect various medical conditions I currently have. Would I have to do it forever? Do my doctors approve? |
|  | [35] Affects on digestion as I gave Crohn's and would be interested in that information. |
|  | [74] How personal medical history, eg, cardiovascular disease, MI- would affect use of Tir. |
|  | [92] I would prefer to see some research on whether it improves, worsens, or has no effect upon the following: Insomnia, IBS, MCAS, Lipedema, Fibromyalgia. |
| **Costs and Insurance Coverage** | [29] Insurance coverage? Out of pocket costs? Other side effects. |
|  | [50] Cost and ease of obtaining |
|  | [64] Cost. Convenience - I'm not going to go to appointments or groups, but am OK doing either on line. |
|  | [73] Would I have to pay for the injections? If yes, how much would that be? |
|  | [83] I would want to know how much the CPAP MONTHLY RENTAL & SUPPLY costs, in addition to the injectable Rx's, would be. I'd ALSO want to know if, once started, I'd be vulnerable I'd be at any given moment to LOSING ACCESS to this effective treatment, ESPECIALLY when seeing SUCCESSFUL results!! |
| **Mechanisms of Action** | [6] The Pharmacokinetics of the drug. I need to understand better the therapeutic window Vs the adverse potential effect. Understand if smaller more frequent doses were tested and if there's a combination that allows less dependence over time. |
|  | [19] Research data on the chemical mechanism. (How does it work?) Side effects (long term, short term), comparison with CPAP and Oral appliances. |
|  | [47] how does it work and the side affects |
|  | [49] How far I might need to travel for visits. Are injections continued indefinitely? How does Terpetizide work within the body? What is it actually doing? |
|  | [72] A good understanding of how the drug works and risks. |
| **Comparative Effectiveness vs Other Options** | [1] what is different between Tirzepatide and WeGovy? why would I change my therapy? |
|  | [8] What's the likelihood of replacing the Cpap with just the Tirzepatide? |
|  | [18] Is Tirzepatide similar to Ozempic regarding to digestion? Would it be necessary to keep taking it for life? Would there be any improvement in the Cpap machine? |
|  | [19] Research data on the chemical mechanism. (How does it work?) Side effects (long term, short term), comparison with CPAP and Oral appliances. |
|  | [51] I would want to know the overall "success rate" for each treatment (Tirzepatide and CPAP) when the patient complies with all of the instructions for use. Ideally the "success rate" would be expressed as a "percentage achievement" of the agreed-upon goals for each treatment. |
| **Special Populations** | [52] I'm not newly diagnosed with sleep apnea and I already use CPAP (since 206) so if this is diagnostic treatment (which my insurance would cover) I would be open to use Tirzepradie as long as it is safe to use and would not interfere with my work (I work as a Flight Attendant) |
|  | [63] More case studies dependent on body types and activity. |
|  | [67] Can you use the Tirzepatide while pregnant? |
|  | [84] Tirzepatide treatment would have to be accepted by Federal Motor Carrier Safety Administration Medical review board for acceptable treatment for commercial motor vehicle operators holding a DOT medical card. |
|  | Currently cpap is the only acceptable treatment (also for Pilots with FAA medical card). Currently "under current and effective treatment " requirements are cpap compliance report. |
| **Trust/Confidence** | [48] If the drug does not have FDA approval I'd like complete information on the drug: how it works, potential side effects, results from previous animal studies. I need to feel confident that what I'm putting into my body is safe - so convince me.<br>[22] how Tirzepatide with affect various medical conditions I currently have. Would I have to do it forever? Do my doctors approve?<br>[24] Would want to consult my doctor for further information.<br>[27] FDA Approval for how long. Success stories or reviews on the medication. Is it a lifetime treatment?<br>[32] I don't trust sleep studies to be accurate. They are unnatural with all the words and tubes taped all over my body. I've had several and NEVER slept as normal. |
| **Logistical Barriers** | [15] I live across from Lake [...]... Monthly trips would not be an option for me as I live on Social Security only. The gas would a terrible cost.<br>[26] One factor for nutritionist in person would be location, I dislike committing to 40+ minute drives to appointments.<br>[49] How far I might need to travel for visits. Are injections continued indefinitely? How does Terpetizide work within the body? What is it actually doing?<br>[38] Yes, I am unable to drive, so seeing a nutritionist once a month in person would be a challenge.<br>[64] Cost. Convenience - I'm not going to go to appointments or groups, but am OK doing either on line. |
| **Quality of Evidence** | [9] how much do we know about this med & side effects or interactions?<br>[30] ADE events reported over time, and study of long term effects would have to be completed and evaluated by peer review prior to adding tirzepatide to my current drug regimen<br>[46] What are the side effects? How many patients were in the study of the side effects over how long a period of time?<br>[54] I'd like to know well the drug itself has been studied thus far, what its efficacy has been and how rigorously any side effect numbers have been verified. I'm interested in any new cpap data as well.<br>[77] I would need detailed information on safety, including percent of participants who stayed with any research study to the end. |
| **Effect and/or Management after Therapy Discontinuation** | [3] Long term effects of both treatments. Also statistics on how quickly /how much weight is gained back after stopping tirzepatide<br>[18] Is Tirzepatide similar to Ozempic regarding to digestion? Would it be necessary to keep taking it for life? Would there be any improvement in the Cpap machine?<br>[22] how Tirzepatide with affect various medical conditions I currently have. Would I have to do it forever? Do my doctors approve?<br>[27] FDA Approval for how long. Success stories or reviews on the medication. Is it a lifetime treatment? |
|  | [45] What happens if you start with Tirzepatide and want to quit taking it? Are there risks to stopping treatment? |
| **Other** | [6] [...] Understand if smaller more frequent doses were tested and if there's a combination that allows less dependence over time. |
|  | [17] The cpap machine is a tether and takes time to clean, fill w water, set up, etc. in addition to the look which is very awkward especially for a woman. It's embarrassing for what ever reason more so for a woman than a man. |
|  | [42] I'm going back to my previous answer. Worrying about power loss and not being able to get good sleep scares me. So, having a shot to help me for a better quality of life sounds awesome. Also, getting your weight controlled by the food you eat is also very important to me. Having a nutritionist sounds awesome once a month. I'm 50/50 on a group, but it may help with accountability if we have good candidates and keep confidentiality. |
|  | [59] I would love to participate in a study if I qualify. |
|  | [57] I am not familiar with Tirzepatide at all so would need to learn more. I do not like using a CPAP machine at all and would love a safe alternative. |

**E-Table 3.**
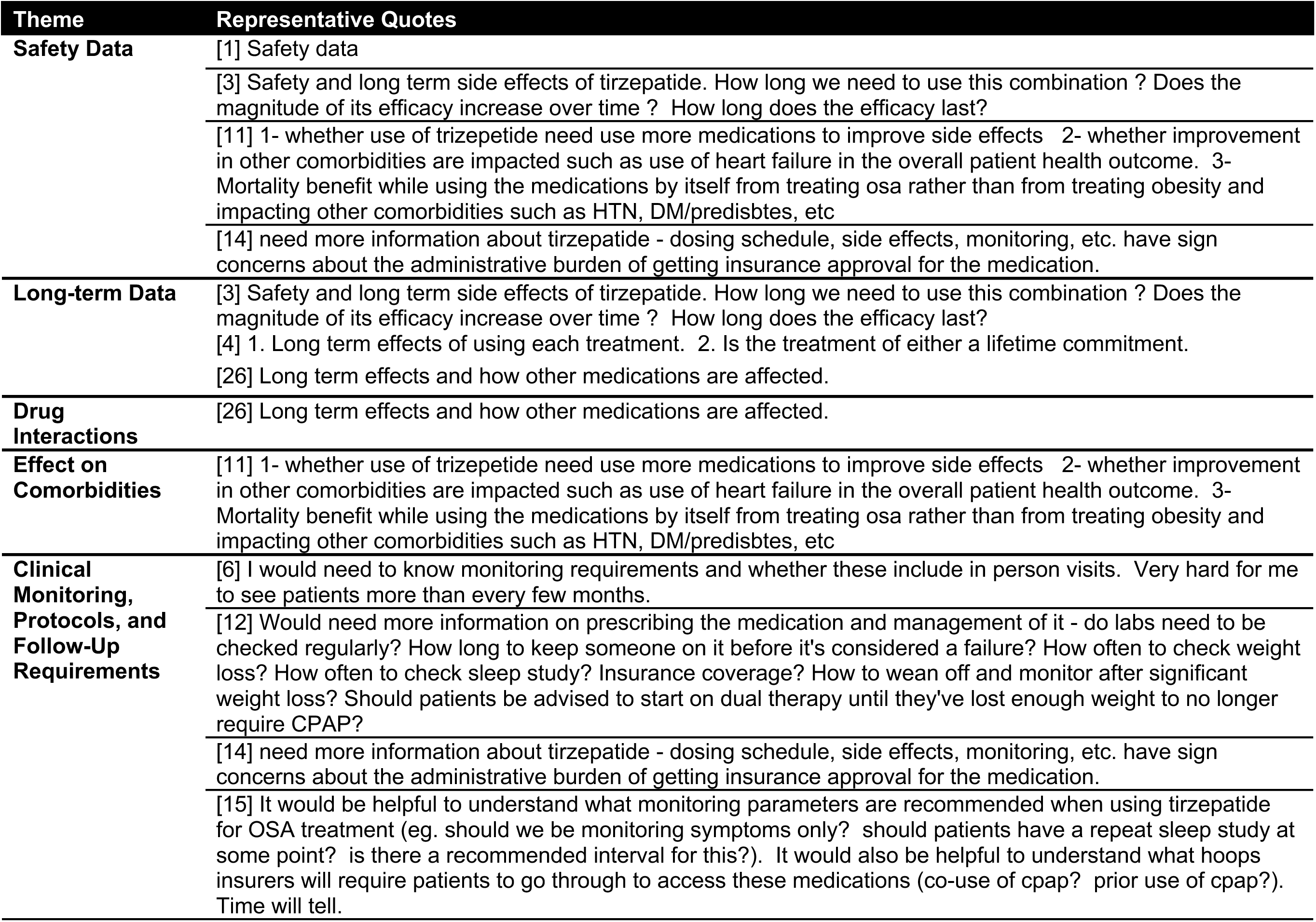
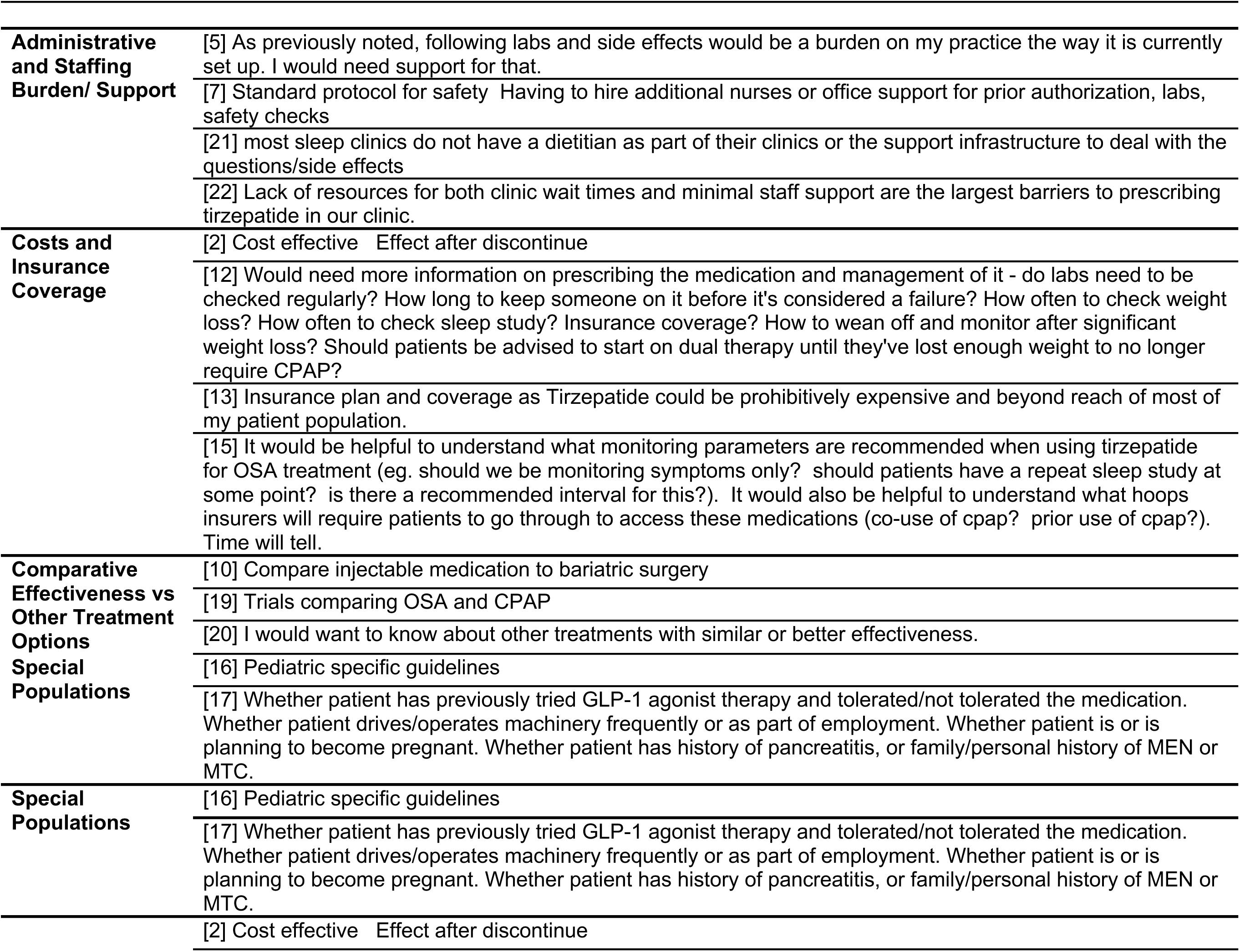
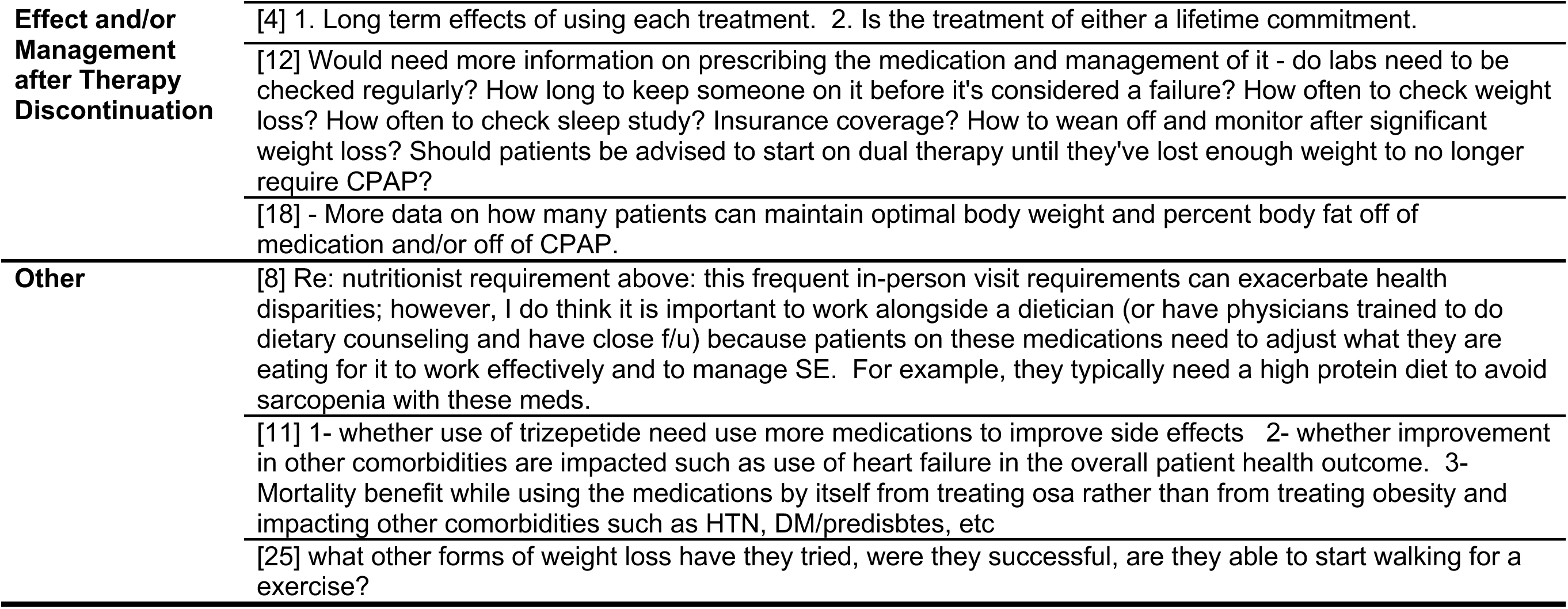
Other Information Needs by Providers: Themes and Representative Quotes. 26 Providers provided free-text comments, some of which related to more than one theme (i.e., 48 related themes).

**E-Table 4.** General Characteristics: Patients from CA vs non-CA. General characteristics were overall similar. CA = California.

| Characteristic | All Patients<br>N = 461<sup>1</sup> | CA Patients<br>N = 404<sup>1</sup> | Non-CA Patients<br>N = 57<sup>1</sup> | P-Value |
| --- | --- | --- | --- | --- |
| **Age** |  |  |  | 0.7 |
| 22-30 years | 15 (3.3%) | 14 (3.5%) | 1 (1.8%) |  |
| 31-40 years | 52 (11%) | 44 (11%) | 8 (14%) |  |
| 41-50 years | 61 (13%) | 50 (12%) | 11 (19%) |  |
| 51-60 years | 71 (15%) | 63 (16%) | 8 (14%) |  |
| 61-70 years | 109 (24%) | 96 (24%) | 13 (23%) |  |
| 71-80 years | 117 (25%) | 103 (25%) | 14 (25%) |  |
| >80 years | 36 (7.8%) | 34 (8.4%) | 2 (3.5%) |  |
| **Gender** |  |  |  | 0.4 |
| Woman | 212 (46%) | 180 (45%) | 32 (56%) |  |
| Man | 244 (53%) | 219 (54%) | 25 (44%) |  |
| Transgender | 1 (0.2%) | 1 (0.2%) | 0 (0%) |  |
| Non-binary/non-conforming | 3 (0.7%) | 3 (0.7%) | 0 (0%) |  |
| PNTS | 1 (0.2%) | 1 (0.2%) | 0 (0%) |  |
| **Race (multiple selections possible; total percent >100%)** |  |  |  | 0.11 |
| American Indian or Alaska Native | 8 (1.7%) | 5 (1.2%) | 3 (5.3%) |  |
| Asian | 31 (6.7%) | 29 (7.2%) | 2 (3.5%) |  |
| Black or African American | 15 (3.3%) | 13 (3.2%) | 2 (3.5%) |  |
| Hispanic or Latino | 44 (9.5%) | 42 (10%) | 2 (3.5%) |  |
| Middle Eastern or North African | 7 (1.5%) | 7 (1.7%) | 0 (0%) |  |
| Native Hawaiian or Other Pacific Islander | 2 (0.4%) | 2 (0.5%) | 0 (0%) |  |
| White | 338 (73%) | 290 (72%) | 48 (84%) |  |
| Other/PNTS | 16 (3.5%) | 16 (4.0%) | 0 (0%) |  |
| **Military Status** |  |  |  | 0.6 |
| Active | 13 (2.8%) | 13 (3.2%) | 0 (0%) |  |
| Veteran | 59 (13%) | 52 (13%) | 7 (12%) |  |
| None/Unknown | 389 (84%) | 339 (84%) | 50 (88%) |  |

<sup>1</sup>n (%); Median (Q1, Q3)
Abbreviations: PNTS Prefer not to say.

**E-Table 5.** Patients Clinical Characteristics and History: Patients from CA vs non-CA. Clinical characteristics and history were overall similar. CA = California.

| Characteristic | All Patients<br>N = 461<sup>1</sup> | CA Patients<br>N = 404<sup>1</sup> | Non-CA Patients<br>N = 57<sup>1</sup> | P-Value |
| --- | --- | --- | --- | --- |
| **Anthropometrics & OSA/Obesity Status** |  |  |  |  |
| **Body Mass Index (BMI), kg/m<sup>2</sup>** | 31 (27, 35) | 30 (27, 35) | 31 (28, 37) | 0.4 |
| Overweight (BMI 25-30 kg/m<sup>2</sup>) | 155 (34%) | 138 (34%) | 17 (30%) | 0.5 |
| Obesity (BMI>30 kg/m<sup>2</sup>) | 247 (54%) | 216 (53%) | 31 (54%) | 0.9 |
| **Likely OSA (Diagnosis or STOP≥2)** | 449 (97%) | 396 (98%) | 53 (93%) | **0.049** |
| OSA Diagnosis, yes vs no/unsure | 398 (86%) | 347 (86%) | 51 (89%) | 0.5 |
| **Likely COBOSA (likely OSA + obesity)** | 245 (53%) | 214 (53%) | 31 (54%) | 0.8 |
| COBOSA (OSA diagnosis + obesity) | 224 (49%) | 193 (48%) | 31 (54%) | 0.3 |
| **Comorbidities, yes vs no/unsure** |  |  |  |  |
| Hypertension | 231 (50%) | 211 (52%) | 20 (35%) | **0.015** |
| Diabetes | 72 (16%) | 63 (16%) | 9 (16%) | >0.9 |
| Mood Disorder | 177 (38%) | 156 (39%) | 21 (37%) | 0.8 |
| **Symptoms (in the past month)** |  |  |  |  |
| **Snoring loudly, yes vs no/unsure** | 263 (57%) | 237 (59%) | 26 (46%) | 0.062 |
| If yes, does snoring disrupt your relationships? | 209 (79%) | 186 (78%) | 23 (88%) | 0.2 |
| **Sleepiness (feeling like may doze off or need to take a nap)** |  |  |  | 0.5 |
| 1+/week | 298 (65%) | 264 (65%) | 34 (60%) |  |
| <1/week | 156 (34%) | 134 (33%) | 22 (39%) |  |
| Unsure/PNTS | 7 (1.5%) | 6 (1.5%) | 1 (1.8%) |  |
| **Fatigue/Lack of Energy** |  |  |  | 0.8 |
| 1+/week | 316 (69%) | 278 (69%) | 38 (67%) |  |
| <1/week | 141 (31%) | 122 (30%) | 19 (33%) |  |
| Unsure/PNTS | 4 (0.9%) | 4 (1.0%) | 0 (0%) |  |
| **Unrefreshing/Poor Quality Sleep** |  |  |  | >0.9 |
| 1+/week | 302 (66%) | 265 (66%) | 37 (65%) |  |
| <1/week | 153 (33%) | 133 (33%) | 20 (35%) |  |
| Unsure/PNTS | 6 (1.3%) | 6 (1.5%) | 0 (0%) |  |
| **Memory Problems** |  |  |  | 0.4 |
| 1+/week | 214 (46%) | 191 (47%) | 23 (40%) |  |
| <1/week | 228 (49%) | 195 (48%) | 33 (58%) |  |
| Unsure/PNTS | 19 (4.1%) | 18 (4.5%) | 1 (1.8%) |  |
| **Therapy for OSA** |  |  |  |  |
| **Ever used treatment prescribed for OSA** | 366 (79%) | 319 (79%) | 47 (82%) | 0.5 |
| **Continuous Positive Airway Pressure (CPAP)** |  |  |  | **0.010** |
| Currently | 285 (78%) | 240 (75%) | 45 (96%) |  |
| In the past | 70 (19%) | 68 (21%) | 2 (4.3%) |  |
| Never | 9 (2.5%) | 9 (2.8%) | 0 (0%) |  |
| Unsure/PNTS | 2 (0.5%) | 2 (0.6%) | 0 (0%) |  |
| **Oral Appliance Therapy (OAT)** |  |  |  | **0.5** |
| Currently | 36 (9.8%) | 33 (10%) | 3 (6.4%) |  |
| In the past | 41 (11%) | 35 (11%) | 6 (13%) |  |
| Never | 281 (77%) | 245 (77%) | 36 (77%) |  |
| Unsure/PNTS | 8 (2.2%) | 6 (1.9%) | 2 (4.3%) |  |
| **Hypoglossal Nerve Stimulation (HNS)** |  |  |  | **0.086** |
| Currently | 6 (1.6%) | 5 (1.6%) | 1 (2.1%) |  |
| In the past | 4 (1.1%) | 3 (0.9%) | 1 (2.1%) |  |
| Never | 348 (95%) | 306 (96%) | 42 (89%) |  |
| Unsure/PNTS | 8 (2.2%) | 5 (1.6%) | 3 (6.4%) |  |
| **Uvulopalatopharyngoplasty (UPPP)** |  |  |  | **0.6** |
| Yes | 24 (6.6%) | 22 (6.9%) | 2 (4.3%) |  |
| No | 332 (91%) | 289 (91%) | 43 (91%) |  |
| Unsure/PNTS | 10 (2.7%) | 8 (2.5%) | 2 (4.3%) |  |
| **Therapy for Overweight/Obesity** |  |  |  |  |
| **Ever tried losing weight on own using diet & exercise** |  |  |  | **0.7** |
| Never | 30 (6.5%) | 28 (6.9%) | 2 (3.5%) |  |
| 1-2 times | 80 (17%) | 69 (17%) | 11 (19%) |  |
| 3-5 times | 88 (19%) | 77 (19%) | 11 (19%) |  |
| >5 times | 250 (54%) | 217 (54%) | 33 (58%) |  |
| Unsure/PNTS | 13 (2.8%) | 13 (3.2%) | 0 (0%) |  |
| **Ever used prescribed therapy for weight loss (e.g., medication or surgery)** | 138 (30%) | 117 (29%) | 21 (37%) | **0.2** |
| **Medical weight loss program (e.g., nutritionist, meal replacement)** |  |  |  | **0.7** |
| Currently | 33 (24%) | 26 (22%) | 7 (33%) |  |
| In the past | 69 (50%) | 59 (50%) | 10 (48%) |  |
| Never | 34 (25%) | 30 (26%) | 4 (19%) |  |
| Unsure/PNTS | 2 (1.4%) | 2 (1.7%) | 0 (0%) |  |
| **Oral weight loss medication** |  |  |  | **0.7** |
| Currently | 20 (14%) | 18 (15%) | 2 (9.5%) |  |
| In the past | 43 (31%) | 38 (32%) | 5 (24%) |  |
| Never | 67 (49%) | 54 (46%) | 13 (62%) |  |
| Unsure/PNTS | 8 (5.8%) | 7 (6.0%) | 1 (4.8%) |  |
| **Injectable weight loss medication** |  |  |  | 0.9 |
| Never | 36 (26%) | 30 (26%) | 6 (29%) |  |
| Unsure/PNTS | 5 (3.6%) | 4 (3.4%) | 1 (4.8%) |  |
| Currently/In the past | 72 (52%) | 62 (53%) | 10 (48%) |  |
| **Tirzepatide** |  |  |  | 0.4 |
| Currently | 31 (32%) | 24 (29%) | 7 (50%) |  |
| In the past | 8 (8.2%) | 7 (8.4%) | 1 (7.1%) |  |
| Never | 57 (59%) | 51 (61%) | 6 (43%) |  |
| Unsure/PNTS | 1 (1.0%) | 1 (1.2%) | 0 (0%) |  |
| **Semaglutide** |  |  |  | 0.2 |
| Currently | 50 (52%) | 46 (55%) | 4 (29%) |  |
| In the past | 21 (22%) | 17 (20%) | 4 (29%) |  |
| Never | 26 (27%) | 20 (24%) | 6 (43%) |  |
| Unsure/PNTS | 0 (0%) | 0 (0%) | 0 (0%) |  |
| **Liraglutide** |  |  |  | >0.9 |
| Currently | 1 (1.0%) | 1 (1.2%) | 0 (0%) |  |
| In the past | 7 (7.2%) | 6 (7.2%) | 1 (7.1%) |  |
| Never | 88 (91%) | 75 (90%) | 13 (93%) |  |
| Unsure/PNTS | 1 (1.0%) | 1 (1.2%) | 0 (0%) |  |
| **Weight loss surgery (e.g., gastric bypass/banding, sleeve gastrectomy)** |  |  |  | 0.9 |
| Yes | 23 (17%) | 19 (16%) | 4 (19%) |  |
| No | 112 (81%) | 95 (81%) | 17 (81%) |  |
| Unsure/PNTS | 3 (2.2%) | 3 (2.6%) | 0 (0%) |  |

<sup>1</sup> n (%); Median (Q1, Q3)

